# Integrative Gene-Centric Analysis of Breast Cancer: High-Confidence Germline Predisposition Genes from Population-Scale Cohorts

**DOI:** 10.1101/2025.09.10.25335496

**Authors:** Reoi Zucker, Shirel Schreiber, Amos Stern, Michal Linial

**Author notes:** **Corresponding authors** Michal Linial.

## Abstract

**Background:** Heritable breast cancer (BC) predisposition is shaped by high-penetrance genes such as BRCA1 and BRCA2, but many moderate- and low-penetrance genes remain uncharacterized. While over 100 independent loci have been reported, their causal genes are often mixed with spurious identifications.

**Methods:** Using large-scale, multi-ethnic genomic data from cohorts including the UK Biobank (UKB) and FinnGen (FG), we refined the catalog of BC predisposition genes by requiring consistency across multiple GWAS in Open Targets (OT) and excluding likely false positives, thereby reaffirming the contribution of established genes such as BRCA1, BRCA2, PALB2, CHEK2, and other DNA repair genes.

**Results:** Our multi-cohort design enabled replication across European ancestry, although transferability to other populations was limited, as shown in the Million Veteran Program (MVP) cohort. By integrating genome, transcriptome-, and proteome-wide association studies (GWAS, TWAS, and PWAS), we identified 38 high-confidence BC predisposition genes, including eight previously reported drivers, 13 genes supported by multiple lines of evidence, and a set of candidates (e.g., APOBEC3A, TNS1, PEX14) that currently lack supporting evidence in the context of heritable BC. PWAS, a gene-based association approach that assess the aggregated effect of coding variants on protein function was applied for the entire UKB population. A dozen candidate genes were identified, most of them were considered within the core gene set, and additional two genes that match recessive inheritance. Overlap with FG data and additional gene-based models further supported the involvement of moderate- and high-penetrance genes such as DNMT3A and ATM.

**Conclusions:** These findings refine the genetic architecture of BC susceptibility, define a core set of predisposition genes, and highlight novel candidates for functional follow-up. The core list of BC susceptibility genes provides biological insights and a framework for gene-based targeted prevention and clinical risk prediction.

## Introduction

Breast cancer (BC) is one of the most common malignancies globally, with its development driven by both genetic predisposition and environmental/lifestyle factors like age, obesity, and hormonal exposure [1-5]. A strong family history of BC is a significant risk factor, highlighting the role of shared genetics and environment [6].

Approximately 5-10% of all BC cases are associated with hereditary genetic mutations [7]. The most well-known high-penetrance tumor suppressor genes, BRCA1 and BRCA2, were identified through family-based linkage studies and are among the most significant BC predisposition genes [8]. The impact of these mutations on cancer risk, particularly for BC, differs between males and females [9]. While BRCA1/2 are clinically important, their overall contribution at the population level is modest. Other high-penetrance genes, such as TP53, RAD51C, RAD51D, and PALB2, also markedly increase lifetime risk [10, 11]. Additionally, polygenic risk scores (PRS) have helped identify moderate-to-low penetrance genes like CHEK2, ATM, PTEN, and PPM1D [12]. The integration of next-generation sequencing (NGS) technologies in clinical oncology now allows for more precise individual risk assessments, especially in specific genetic subpopulations [13-15].

Despite the growing body of genetic data, there is no consensus on a definitive set of genes that govern BC genetic risk, and their general applicability across diverse populations remains unclear. While some candidate predisposition genes are included in testing panels, treatment and therapy strategies based on these findings are not yet conclusive [16-18]. The field has been shaped by numerous associated variants from Genome-Wide Association Studies (GWAS). To date, about 200 independent loci have been linked to BC susceptibility, though most were identified at sub-genome-wide significance thresholds (e.g., p <1.0e-06). These loci generally have small or negligible effect sizes (odds ratios <|1.1|), but their cumulative impact has been used in Polygenic Risk Scores (PRS) for risk stratification at the population level [19], and specifically for carriers of high-penetrance variants [20]. The importance of adding PRS for refining clinical stratification was confirmed [21].

Cross-cohort replication studies have further refined our understanding of population-specific risk architectures. For example, the Finnish genetic project (FinnGen, FG) [22] has uncovered rare, population-enriched variants in known BC genes (BRCA1/2, RAD51C, BARD1) due to founder effects [23]. Similar founder mutations in CHEK2 (1100delC) and PALB2 have been identified, with similar genes found in Korean women [24]. These large-scale genetic resources validate known risk genes and identify novel, population-specific variants, underscoring the roles of evolutionary forces like migration and genetic drift [25]. A notable example is a large BRCA1 deletion (exons 9–12), a founder mutation among women of Mexican descent that contributes to a high rate of hereditary BC [26]. Three specific founder frameshift mutations, BRCA1 (185delAG, 5382insC) and BRCA2 (6174delT), are approximately 10-fold more prevalent in certain populations than in the general population. In East Asia, BRCA1 (c.5470_5477del) and BRCA2 (c.3109C>T) are recurrent in the Chinese Han population, while BRCA1, L63X and BRCA2 7708C>T are prominent in isolated Japanese groups. CHEK2 mutations also show a higher prevalence in East Asian populations compared to Europeans. Among Icelanders, BRCA2 999del5 is a population-specific stop-gain mutation found in about 1% of individuals. The BRCA1 943ins10 mutation is more frequent in African American cohorts, which also exhibit higher rates of triple-negative BC. High consanguinity rates also drive the occurrence of pathogenic mutations, as seen with the unusual prevalence of BRCA1 c.68_69delAG in Palestinian women. These patterns illustrate that inherited BC risk is shaped by both individual gene effects and broader population genetic structures.

The UK Biobank (UKB) is a large resource of 500,000 participants, predominantly of European descent [27]. In the UKB, pathogenic BRCA1/2 variants are found in less than 0.5% of women, and most BC cases occur in individuals without any known pathogenic mutations. The hereditary component of triple-negative BC (TNBC), which has a poor prognosis, shares features with BRCA1-deficient tumors. Approximately 70 loci have been identified through GWAS and large-scale replication studies [28]. A conservative list of about 20 predisposition genes can be categorized by their functions in cell cycle, DNA repair, adhesion, and endothelial physiology [29]. Understanding the distribution of these mutations across different ethnic and geographic groups is crucial for effective screening, genetic counseling, and equitable access to precision medicine [25, 30].

This study addresses several understudied aspects of BC predisposition genetics. Our analysis focuses on detecting susceptibility genes rather than causal variants. First, we revisit the list of BC-GWAS credible genes by leveraging external resources like the Open Targets (OT) gene-association platform [31] to prioritize genes based on genetic association and non-genetic support. Our analysis covers a range of variants, from high-effect ones like BRCA2 to common variants with smaller effects. Second, we re-examined large biobank samples (UKB, FG, and MVP) to assess the benefit of finding gene-level coherence and agreement to improve the discovery rate of genes with low and modest effect sizes. To overcome inconsistencies in variant mapping often seen in meta-analyses, we used credible gene lists from FinnGen GWAS of a refined cohort of BC patients who were not affected by other cancer types. Finally, we reanalyzed the data using multiple genetic association methods, including PWAS for coding region variants, TWAS for variants that alter gene expression, and several sets of GWAS. This led to a confident core list of BC susceptibility genes that can be used for further research and improving the accuracy of BC risk management at both the population and individual levels.

## Materials and Methods

### Biobank data processing

The UK Biobank (UKB) is a population-scale resource that integrates deep phenotypic, genotypic and lifestyle information on roughly 500 000 volunteers aged 40-69 recruited across the United Kingdom between 2006 and 2010 with follow-up and ongoing update of diagnosis (2024-Q3 data release). For genetic analysis, we removed related individuals by keeping randomly chosen participant per kinship cluster (Zucker 2023). In the PWAS results, the samples were further restricted to genetically inferred Europeans (UKB data-coding 1, 1001, 1002, 1003 and field 21000). A complementary, whole-cohort view retains the∼ 78,000 participants labelled “non-white-British”. We adhere to the definition from PheCode 174.11 (Supplementary **Fig. S1**). based on mapping to ICD-10 (International Statistical Classification of Diseases and Related Health Problems,10th Revision) indexed C50; UKB field 41270 [32].

FinnGen Freeze 12 (November 2024, FG12) provides >21.2 M variants (deep sequencing enriched) from >500,000 Finnish individuals (median age 63 y, 56.4 % women). Standard quality filters removed variants with >2 % missingness, HWE P <1e-06 or MAC <3. All FG12 analyses adjusted for sex, age, 10 PCs, chip version and batch. We analysed the results from the cohort of C3_BREAST_EXALLC with 24,270 cases and 222,078 controls. In addition, we tested subtypes of breast cancer (C3) marked as C3_BREAST-ERNEG_EXALLC (9,724 cases) and C3_BREAST-ERPLUS_EXALLC (14,540 cases). There are 221,705 controls in FG12. We reanalyzed the GWAS fine-mapping with Bayes factors, yielding high-confidence variants. We also analyzed the coding-variant subset and the PheWAS interrogation activated by Ristey R13 query interface [22]. FG12 was considered a disjoint cohort, and used for replication of gene discovery, and increasing confidence. We considered only the credible set (CS). Specifically, each variant is associated with a value that captures the likelihood to be causal for a phenotype. The analysis reports as credible set (CS) where confidence set (CS)) is used as a measure for fine-mapping analyses to assess the probability that a given genetic variant is causal for the disease.

### GWAS, coding-gene GWAS and summary statistics analyses

The data from the UKB was analyzed as described before [33]. We performed GWAS in two versions, the routine GWAS and the gene-length GWAS (cgGWAS). Among the 487,409 participants with inpatient records (field 41,270, September 2024), 16,952 carried at least one C50 code, 64% were diagnosed hospital admission. The GWAS-ready C50 comprised 16,952 cases and 409,651 controls with genotyping imputed data. GWAS was based on the genotyping data with approximately 820k chosen genetic variations available for every participant. We employed a standard PLINK procedure for the analysis. The genotyping information, are based on the UKB Axiome Array. We applied UKB imputation protocol and the count 97,013,422 variants. For the imputed variants, we computed the probabilistic expectations for the alternative alleles [34]. We have conduct analysis that is based on coding gene length (cgGWAS) using the MAF threshold >0.001, Hardy-Weinberg equilibrium (HWE), with p-value of 1e-06, and variant calling genotyping coverage at ≥90%. In total, we examined >10 M variants. We additionally added sex, year of birth, and the initial six principal components (PCs) as covariates to consider population structure (total 8 covariates).

We used the summary statistics reported by Open Targets (OT) platform (release date: 9/2024; Ghoussaini, 2021 #34}) to collect the current knowledge on BC GWAS results The BC used for the OT is a compilation from multiple resource including EFO: MONDO_0007254, UMLS: C0006142, NCIt: C9335. This broad definition includes the cases of Malignant Breast Phyllodes Tumor, Breast carcinoma, Breast lymphoma, Malignant breast melanoma and Breast sarcoma as described in OT. Each gene is listed with OT global score (range 0-1.0). Among these genes, 1825 genes are supported by any evidence and 217 were associated with genetic-associated (GA) scores based on large-scale independent GWAS summary statistics [35]. Each locus is associated with genes by to their location and based on their association according to linkage disequilibrium (LD).

### Gene-level effect scores across the human proteome

The PWAS framework [34] posits that causal coding variants affect phenotypes by altering the biochemical activity of their corresponding proteins. To quantify this, the pretrained ML model FIRM [34] estimates the impact of variants for each protein across the proteome. Validated against ClinVar pathogenic variants, FIRM demonstrated strong performance (AUC = 90%, accuracy = 82.7%) [34]. Variant effect scores are scaled from 0 (loss of function. LoF) to 1 (synonymous changes, no effect) [34]. From ∼97 million variants, 639,323 coding and splicing variants spanning 18,053 protein-coding genes were analyzed [36]. To estimate individual-level protein damage, per-variant predictions are aggregated at the gene level under recessive, dominant, and hybrid inheritance models. On average, each gene-level effect score reflects contributions from ∼35.4 nonsense and missense variants.

To determine the effect size of a gene, we applied a measure of Cohen’s d values. Cohen’s d, also known as standardized mean difference, measures the difference between two means divided by a standard deviation (SD) for the data. In this study, Cohen’s d is the (normalized) difference in mean gene effect scores between cases and controls (calculated independently for both dominant and recessive effect scores). The variant association and effect size by routine GWAS were calculated by PLINK 2.0 default logistic regression. The calculated z scores specify the effect size and its directionality. Note that in GWAS, a positive z score indicates a positive correlation between hypothyroidism and the number of alternative alleles, thereby indicating a risk variant. In PWAS, positive values indicate a positive correlation with the gene effect scores, whose higher values mean less functional damage. Thus, negative values are indicative of protective variants in GWAS versus risk genes in PWAS.

### Transcriptome-wide association study (TWAS) analysis

We applied transcriptome-wide association studies (TWAS) [13], which integrate publicly available GWAS summary statistics with transcriptomic prediction models. Specifically, we used TWAS-Atlas built on European ancestry data. TWAS employs a framework which imputes gene expression across 44 human tissues simultaneously using a multi-task learning strategy [37]. Gene–disease associations is determined by combining association scores from single tissues into a cross-tissue test. TWAS offers a broad resource of models linking genomic loci to disease risk [13]. TWAS frequently yields an expanded list of candidate genes, as mapping of a locus to gene is often reports on >10 genes per locus,. In this study, we did not examine coherence across different TWAS models.

### External comparative analyses

To benchmark our findings, we compared them with results from gene-based genotype–phenotype associations available through ExPheWAS [38]. ExPheWAS is a phenome-wide association study (PheWAS) platform that reports associations for 26,616 genes (including long non-coding RNAs), across a large number of phenotypes in ∼410,000 UKB participants. Additionally, we reviewed GWAS summary statistics from the Global Biobank Engine (GBE), which compiles data from >750,000 individuals across the UKB, the Million Veterans Program (MVP), and BioBank Japan. The phenotype studied is based on 16,136 cases of BC (indexed cancer1002) [39]. The GBE analysis applied consistent disease definitions and Bayesian methods to evaluate genetic effects. Notably, log10(Bayes Factor) ≥2 was considered to provide moderate evidence supporting the reliability of a gene or variant’s effect on a given phenotype [39].

### Bioinformatics tools

For functional enrichment of annotations and pathways, we applied the Gene2Func function of FUMA-GWAS using default parameters and a set of genes as input [40]. All values are reported by their adjusted p-values, using the human gene coding proteome as background. The enrichment analysis is based on statistical methods considering multiple hypotheses. The report relies on the significant functional annotations where each finding is associated with a weighted combination of p-value and z-score (combined score).

### Resource and availability

The analysis shown in this study are supported by Supplementary materials (Fig. S1-S5 and Supplementary Tables S1-S10). GWAS (with 10 covariates) and PWAS (with 10 covariates) performed in this study for ICD10: C50 (for female only, and both sexes). The results from these runs are shared in Zenodo data repository (10.5281/zenodo.17086606).

For summary statistics GWAS comparison we utilize the compilation from Open Targets (OT) platform https://platform.opentargets.org/disease/MONDO_0007254. Exclusion and inclusion rules per outcomes and phenotypes from FinnGen are found in https://risteys.finngen.fi/endpoints. GBE analysis used the phenotype names https://biobankengine.stanford.edu/RIVAS_HG19 (index cancer1002). The UKB analysis from Neals lab summary statistics (2418 phenotypes) based on 337,159 participants, resulted in 113 variants is reported in https://pheweb.org/UKB-Neale/pheno/20001_1002#qq. The computational pipeline for processing the UKB data used in this work is an open-source project available at https://github.com/nadavbra/ukbb_parser. FIRM model and prediction of variant-centric effect score (https://github.com/nadavbra/firm). The PWAS is available in https://github.com/nadavbra/pwas. PWAS analysis on Europeans is presented in PWAS Hub (https://pwas.huji.ac.il/). Lavaa plot visualization application generated genetic volcano plots were available in PheWAS browser lined to FinnGen https://mvp-ukbb.finngen.fi/variant. ExPhWAS browser is available in https://exphewas.ca/v1/docs/browser and allows searching for the breast cancer phenotype of PheCode 174.11 (female only).

## Results

### Integrative framework for breast cancer (BC) gene identification and validation

The goal of this study is to compile a set of genes that can be confidently assigned as a predisposition breast cancer (BC) core set. The genetics of BC has been extensively studied; however, results from complementary association studies and gene-based methods remain inconsistent. As a result, a gap persists between BC genetics, risk assessment, and clinical application. Here, we present a framework that seeks coherence across large population studies, validates BC core genes, and proposes clinical utility.

**Fig. 1A** illustrates the multi-layered approach used in this study to identify and validate core BC genes. The main strategies include several stages (**Fig. 1A**, left to right). Firstly, applying filtering protocols using the Open Targets (OT) platform based on results from multiple GWAS and their credible sets (CS). Then, comparing germline results to somatic BC driver gene lists to test the contribution of established cancer genes to BC predisposition. We further leveraged classical and coding-gene (cg) GWAS from the UKB cohort to assess gene-level coherence and replication in an independent FinnGen (FG) cohort with their CS gene lists. For estimating population-origin coherence and variant effect sizes, the Finnish GWAS results (FG12) with summary statistics from the Million Veteran Program (MVP) were partition to includes European- and African-ancestry BC cohorts. Lastly, we integrated complementary protocols from OT-filtered genes identified through GWAS, and other genome-wide association methods such as TWAS (based on eQTL), and PWAS (based on predicted protein function impact). **Fig. 1B** presents the converged results from these approaches, yielding the core BC gene list. These genes are further validated using external evidence, including clinical panels, FG coding, BC subtype data, somatic drivers, gene function, and published literature. Finally, this framework demonstrates the clinical utility of evaluating population stratification in relation to BC risk.

**Figure 1.**
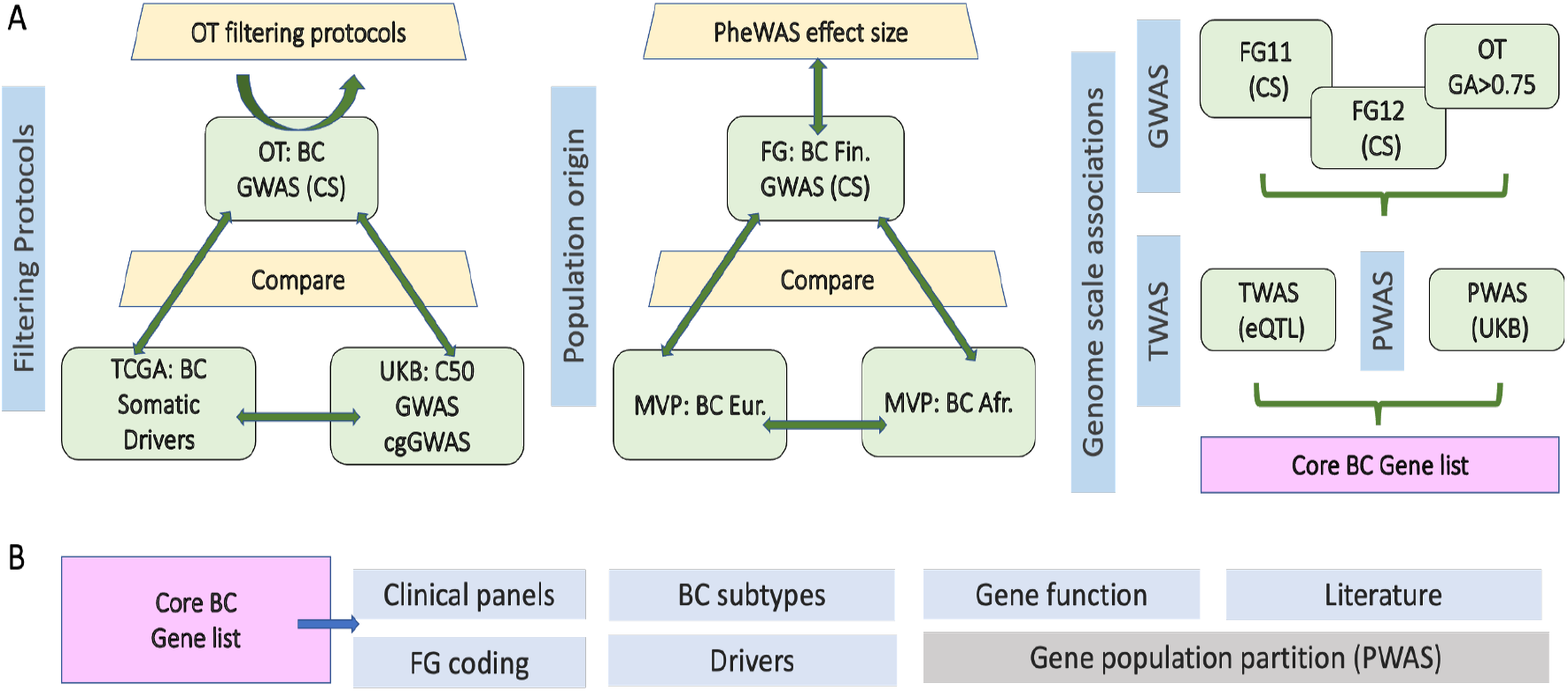
The outline of the framework presenting for BC core gene set and validation. **(A)** The protocol used for filtering the OT gene list based on multiple BC GWAS. Testing GWAS genes with BC driver genes from somatic data. The diversity of the population origin in illustrated by NVP data from european and african origin. A complementary set of GWAS, PWAS and TWAs were combined to present a core BC gene list. **(B)** Validation through external resources, The utility of population partition is tested for PWAS results. BC, breast cancer; FG, FinnGen; OT, Open targets; UKB, UK biobank; CS, credible variant set mapped to genes.

### Germline risk genes for breast cancer (BC) and somatic driver genes are largely distinct

We investigated the relationship between germline risk genes and somatic driver genes for BC. Genome-Wide Association Study (GWAS) results for BC were compiled using the Open Targets (OT) platform, which assigns a genetic association (GA) score to each gene (**Fig. 2A**). Out of 638 genes with a GA score, we focused on the subset with GA score ≥0.5. As anticipated, CHEK2 and BRCA2 were ranked at the top. However, approximately 100 genes also demonstrated high GA scores (>0.7), suggesting a broader genetic landscape for BC predisposition than previously assumed (Supplemental **Table S1**).

**Figure 2.**
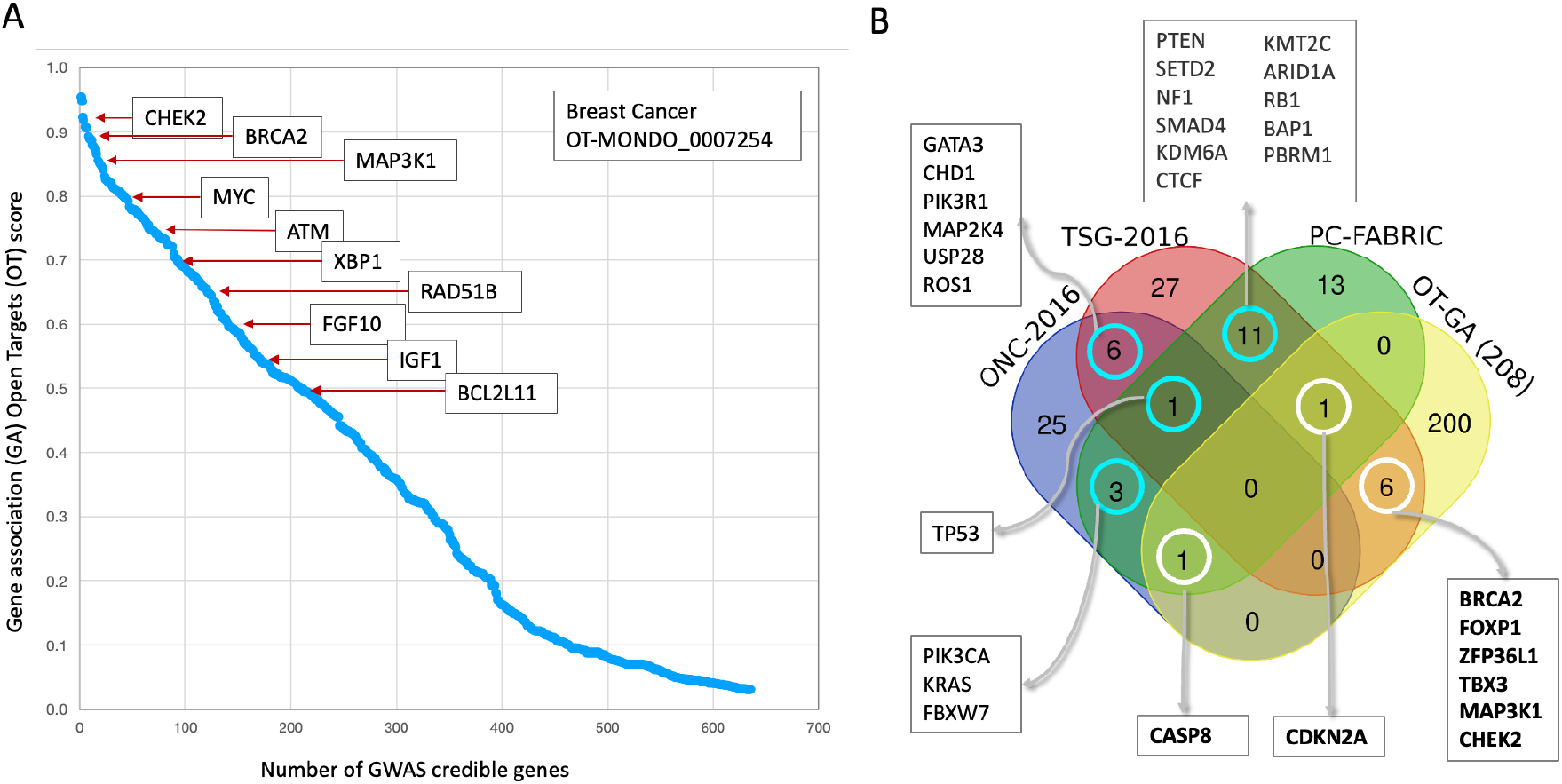
GWAS BC-credible genes from OT platform and overlap with driver genes. **(A)** A plot showing the distribution of genetic association (GA) scores for 638 genes identified by the Open Targets (OT) platform, with a few selected genes that have a high GA score ≥ 0.5 (total 208 genes). **(B)** A Venn diagram illustrating the overlap between three gene sets. A list of 208 BC-credible genes from the OT platform (with GA ≥ 0.5), a list of driver genes from primary BC samples, partitioned to oncogenes (ONC, 36 genes) and tumor suppressor genes (52 genes, TSG) [30], and a pan-cancer 30 driver genes from FABRIC [41]. The diagram highlights the minimal overlap, with 8 genes from the OT list also identified. The diagram also shows the overlap between the FABRIC list and the oncogene (ONC) and tumor suppressor gene (TSG) lists from primary BC samples, noting that 6 genes appear in both the ONC and TSG categories, indicating their context-dependent function.

To determine if BC predisposition genes share function with cancer drivers, we compared the list of 208 OT genes with a GA score ≥0.5, with established BC-derived driver gene lists. We used two primary resources for driver genes: (i) a comprehensive analysis of 2,433 BC samples that identified oncogenes (ONC) and tumor suppressor genes (TSG) [30], and FABRIC, a machine learning method developed to detect pan-cancer drivers from TCGA data [33]. FABRIC identifies cancer genes under positive selection by comparing the impact of variants on protein function against an expected baseline [41].

Our analysis showed minimal overlap between the GWAS-based predisposition genes from the OT platform and the somatic driver gene lists (**Fig. 2B**). Although there was a statistically significant overlap with the TSG list (hypergeometric p-value <0.01), there was no enrichment of driver genes among the GWAS derived BC predisposition genes. Our findings confirm that only a small subset of drivers is detectable from large-scale cohorts, with 200 of the 208 genes showing no evidence of being a cancer driver gene. The overlapping TSGs were BRCA2, FOXP1, ZFP36L1, TBX3, MAP3K1, and CHEK2. Additionally, CASP8 and CDKN2A were identified through their overlap with FABRIC and assigned as ONC and TSG gene, respectively. Notably, a polymorphism in CASP8 (rs1045485) has been shown to significantly reduce BC risk in certain European and Chinese populations [42, 43]. CDKN2A is a key gene in the cell cycle which was initially associated with melanoma-prone individuals. However, it was shown to increase the prevalence of a broad spectrum of cancer types [44].

We concluded that while GWAS-identified gene lists are likely to be noisy due to false discovery, the small set of genes with germline mutations that also function as somatic BC drivers are strong candidates for BC predisposition.

### Prioritizing candidate genes from the abundance of GWAS false positive genes

To prioritize the list of potential BC susceptibility genes, we focused on 208 candidates with GA score ≥0.5. We then looked for genes within this subset that had a higher global score relative to their GA score reported by the OT platform. This approach allowed us to select genes supported by independent evidence, such as data from scientific literature, animal models, burden test, ClinVar and genetic constraints. Using this protocol, we identified 10 such genes **(Table 1)**. While some, like ATM, BRCA2, and RAD51B, were already well-known BC predisposition genes, others were not known. Some had been previously reported as driver candidates in BC tumors but were not considered germline BC predisposition genes. Comparing the OT global score relative to GA score per gene, demonstrates that prioritizing genes from GWAS can be used for removing false positive and ranking genes based on the strength of independent, external evidence.

**Table 1.** Top genes that were signified by stronger evidence beyond genetic association.

| Gene | Main feature / Function | Relevance to BC predisposition | Role, [penet.]<sup>a</sup> | Population<sup>b</sup> |
| --- | --- | --- | --- | --- |
| CCNE1 | Cell cycle: G1–S transition | Amplified in aggressive BC | BC driver | No |
| C11orf65 | Mitochondrial-related gene | ? | ? | No |
| PTPN11 | Phosphatase in RAS/MAPK signaling | Somatic mutations in BC tumors | ? | No |
| RAD51B | DNA repair, homologous recombination | Low-penetrance gene | [Low] | European & Asian |
| BRCA2 | DNA repair, homologous recombination | Strong association, hereditary BC | [High] | Ashkenazi Jews, Icelandic, French Canadian |
| DNMT3A | DNA methylation, epigenetic regulation | Common in blood cancers | ? | No |
| ATM | DNA damage response; cell cycle checkpoints | Moderate-risk gene | [High-Mod] | European |
| ERBB4 | EGFR family TK receptor | ? | ? | No |
| PALB2 | BRCA2 binding partner; DNA repair | Strongly linked to hereditary BC | [High-Mod] | Finnish, French Canadian, Polish |
| ESR1 | Estrogen receptor; hormone signaling | Common in ER-positive BC | BC driver | No |

<sup>a</sup> Penet., gene penetrance category. Mod, moderate; ?, no consensus for the role of penetrance in BC context.
<sup>b</sup> Population specificity with founder variants.

To further improve our discovery rate, we inspected the subset of 208 genes with high GA scores (≥0.5) that had a lower global OT score. We analyzed the set of gene by their functional enrichment test using FUMA-GWAS. From an initial list of 26 genes (Supplementary **Table S1**), we identified eight that were significantly enriched in at least two of the four tested gene sets: PEX14, WDR43, CCDC170, DNAJC1, ZNF365, CCDC88C, TOX3, and FTO (**Table 2**). Out of all GWAS experiments reported by FUMA-GWAS, all results (at p-value <1e-04) resulted in a strong enrichment for phenotypes related to BC and breast physiology.

**Table 2.**
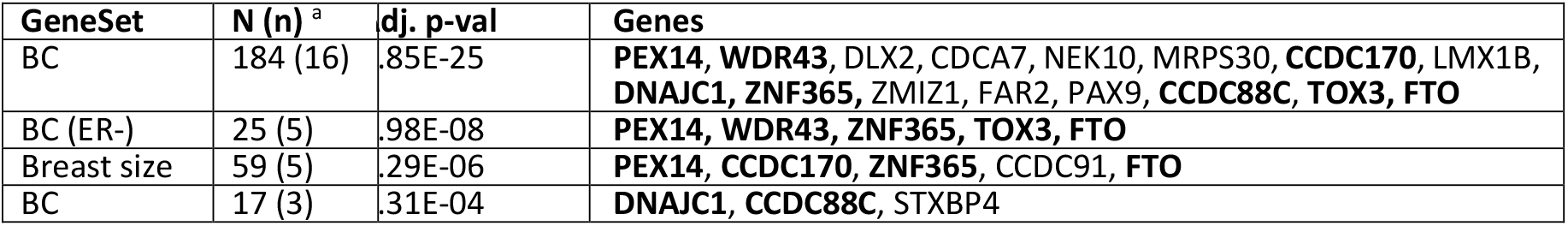
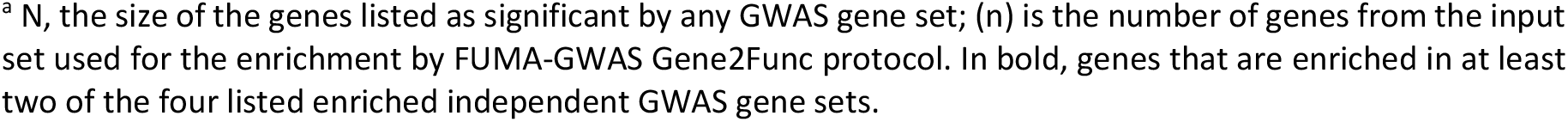
Enrichment test (FUMA-GWAS) for genes signified by markedly reduced OT global scores relative to their GA score.

| GeneSet | N (n)<sup>a</sup> | dj. p-val | Genes |
| --- | --- | --- | --- |
| BC | 184 (16) | .85E-25 | **PEX14, WDR43, DLX2, CDCA7, NEK10, MRPS30, CCDC170, LMX1B, DNAJC1, ZNF365, ZMIZ1, FAR2, PAX9, CCDC88C, TOX3, FTO** |
| BC (ER-) | 25 (5) | .98E-08 | **PEX14, WDR43, ZNF365, TOX3, FTO** |
| Breast size | 59 (5) | .29E-06 | **PEX14, CCDC170, ZNF365, CCDC91, FTO** |
| BC | 17 (3) | .31E-04 | **DNAJC1, CCDC88C, STXBP4** |

<sup>a</sup> N, the size of the genes listed as significant by any GWAS gene set; (n) is the number of genes from the input set used for the enrichment by FUMA-GWAS Gene2Func protocol. In bold, genes that are enriched in at least two of the four listed enriched independent GWAS gene sets.

Manual inspection of genes with a GA score > global OT score, coupled with solid statistics across multiple GWAS (**Table 2**), confirmed the relevance of several BC predisposition candidates. PEX14, a gene involved in peroxisome biogenesis, was recently implicated in BC predisposition by a family-based study [45]. CCDC170 that is located adjacent to the ESR1 gene, shows a consistent association with triple-negative BC (TNBC) across multiple GWAS. Fusion events and specific risk alleles have been linked to aggressive BC subtypes, with a particularly strong effect in the East Asian population [46]. Variants in ZNF365 have been repeatedly associated with BC risk, especially in individuals with a BRCA2 mutation [47]. TOX3 is a known low-penetrance BC susceptibility gene, with common risk variants found in both European and Asian cohorts. Lastly, WDR43 was validated as an ER-negative susceptibility locus through meta-analysis and further confirmed by functional and eQTL studies [48].

We confirmed that evidence-based consideration is valuable for validating known and proposing new candidates as BC predisposition genes (**Table 1**). We also proposed that despite a lack of strong non-genetic evidence, valid candidates display a consistent enrichment in BC-related phenotypes from independent GWAS findings (**Table 2**). Although further confirmation is needed, this simple ranking protocol improves the discovery rate while removing a large fraction of false positives.

### Germline risk genetics for BC is sensitive to population origin

We performed routine GWAS on the imputed variants from UKB. We have used breast cancer diagnosis (ICD-10: C50) and included 10 covariates (sex, age, and top 8 strongest PCs to account for major population structure). There are 16,952 cases and 409,651 controls (among them, 216,254 females). There are only 139 BC cases among the male cohort. We performed the GWAS analysis for both sexes and identified 2,727 variants that met the whole genome GWAS threshold of p-value <5e-08. Among them, only 31% of the variants are associated with a gene (i.e., a successful variant-to-gene mapping), the rest are intergenic and will be further discussed. All together, we report on 137 different genes (Supplementary **Table S2**). Based on the assumption that among the GWAS there are many false positives, we filtered out the list by the following criteria: (i) We only considered variants with minor allele frequency (MAF) of >1% to avoid statistical biases. This removed 15% of the significant variants. (ii) We concentrate on genes that are supported by at least 2 variants with the vast majority of the variants of a gene shares the same directionality. A filter for multiple occurrences of variants for the same gene further confirms the gene relevance, its effect size (measured by the Odds ratio, OR) and coherence in directionality. There are 32 such genes. The other 105 genes that were supported by a single statistically significant variant and therefore were excluded. Notably, extreme cases were detected with many variants per gene as for NEK10 (264 variants), FTO (82 variants), TOX3 (67 variants), EGFR2 (59 variants), MAP3K1 (50 variants) and MRTFA (45 variants). Not only these genes were validated, but their effect size was substantial. For example, the OR of EGFR2 and TOX3 are approximately 1.28 and 1.24, respectively.

To enhance the interpretability, we performed restricted GWAS while limiting the variants to be located in gene length of coding genes (called cgGWAS). There are 531,246 variants that met the coordinated of coding genes (18,053 genes and the included imputed variants), among them 488,783 variants were mapped in the female cohort. Only 11 variants complied with an exome-scale threshold of 5e-07 with two genes that were represented with 2 variants each (CUL7 and CCDC170; Supplementary **Table S3**). **Fig. 3A** shows the Manhattan plot for the female cohort for IDC-10 C50, indicating the 9 significant genes. The vast majority of the statistically significant variants (>95%) are rare and ultra-rare (AF<0.001). The variants associated with CUL7, LSM8 and ACO2 (**Fig. 3A**) are based on ultra rare variants (AF<0.001). **Fig. 3B** shows the quantile-quantile (QQ) plot that compares the distribution of observed and expected distribution of the p-values under the null hypothesis (i.e., no associations). The systematic inflation of test statistics (γ = 1.073) confirms the insignificant inflation across all datapoints. While the GWAS included covariates, we concluded that the GWAS results are not stable enough to provide a solid and reliable BC predisposition gene list.

**Figure 3.**
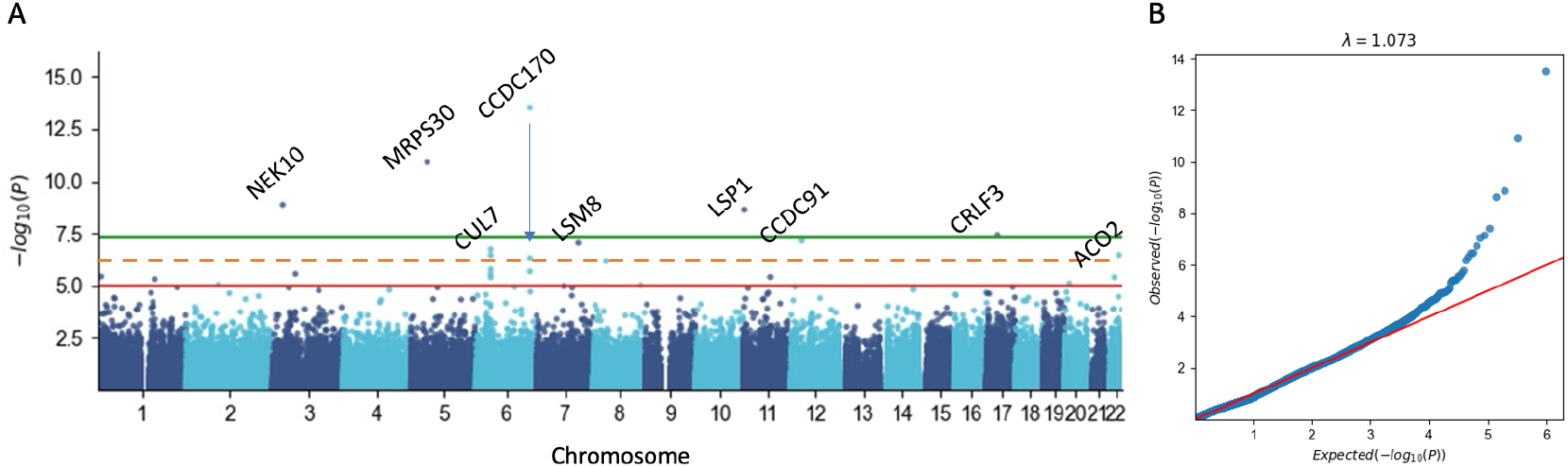
The GWAS results for predisposition in BC. **(A)** Manhattan plots for ICD-10: C50. The plots show the significance of all the variants and genes tested with cgGWAS (see Methods). Among them 488,783 variants in the female cohort, only 11 variants mapped with an exome-scale threshold of 5e-07 (Supplementary **Table S3**, listed all variants with p-value <5e-06), with two genes represented by 2 significant variants (Cul7 and CCDC170). The gene names are indicated by the genes meeting the dashed red line threshold at -log10(P) of 6.3 representing p-value <5e-07. The red and green lines indicate a threshold of 1e-05 and 5e-08, respectively. Note that ChrX and ChrY were not included in the analysis. (**B**) QQ plot comparing the distribution of observed and expected distribution log of the calculated p-values of the null hypothesis (i.e., no associations). Inflation of test statistics (γ) confirms the minimal inflation.

We then tested how using different populations can provide insight into the genetic mechanisms driving BC risk. We used a cohort from FinnGen freeze 12 (FG12) dataset which defines BC cases as malignant neoplasm of breast while excluding all other cancer occurrences from the control group (Supplemental **Fig. S2**). This compilation included 24,978 participants (abbreviated C3_BREAST_EXALLC).

We then analyzed the coherence of associated variants by comparing the FG12 cohort to the Million Veteran Program (MVP), which was split into European and African ancestry groups, as well as the UK Biobank (UKB) cohort which doesn’t overlap with FG. **Fig. 4** compares the allele frequencies (AF) and effect sizes of 137 variants identified in the FG12 meta-analysis cohort to those in the MVP European and African groups. A notable finding was that 26% of the variants from FG12 were missing from the MVP’s European-ancestry group. This is consistent with the importance of founder effects and population bottlenecks in the Finnish population, and the noisy GWAS results.

**Figure 4.**
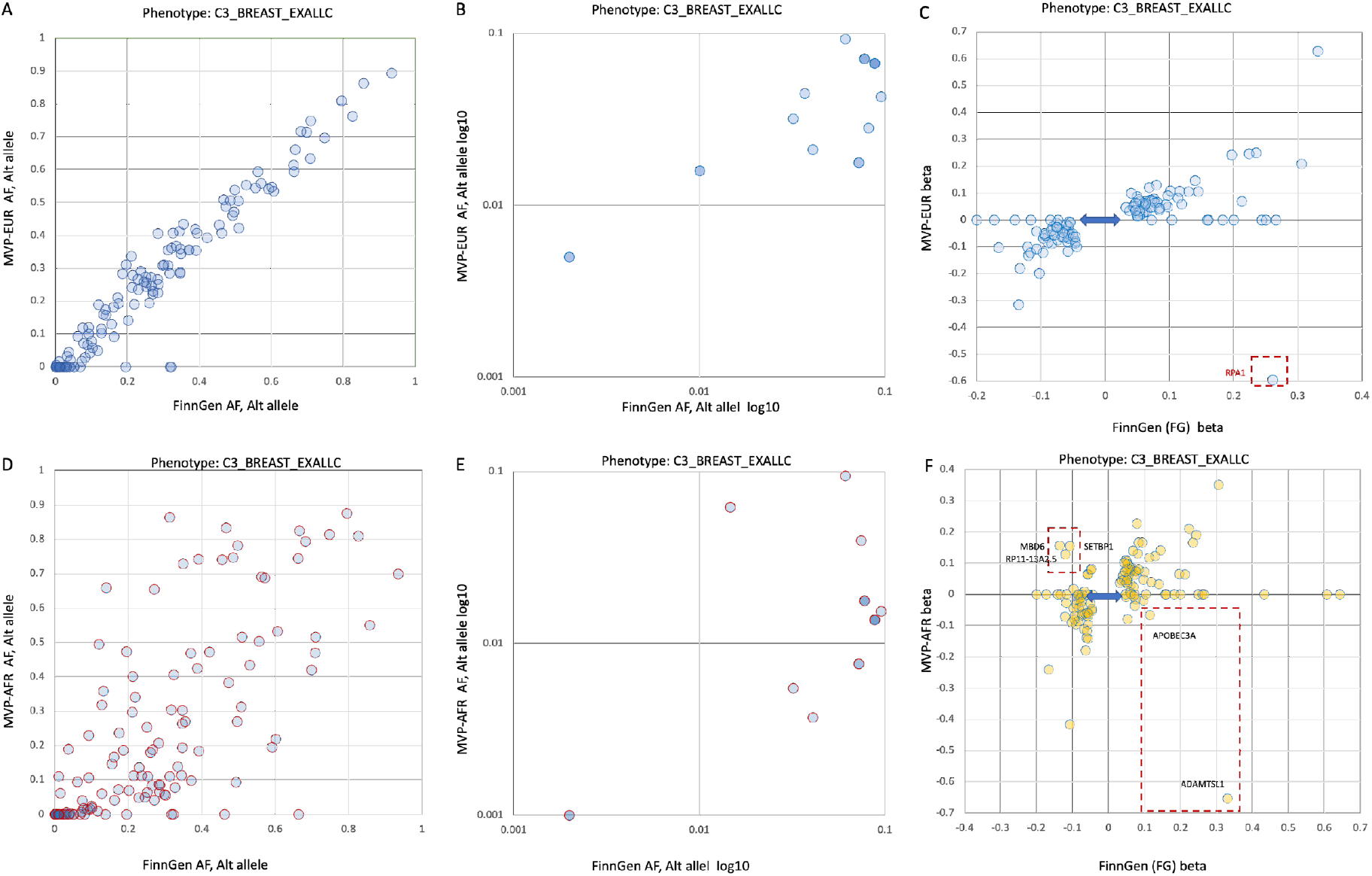
Comparisons of BC-associated variants from B3_BREAST_EXALLC meta-analysis between the FinnGen (FG) cohort and the MVP cohorts. **(A)** Scatter plot of the allele frequencies (AF) between individuals in FG12 and MVP participants of European ancestry (MVP-EUR). (**B)** Scatter plot of the AF between individuals in FG and MVP participants of European ancestry (MVP-EUR) for the AF range of 10% to 0.1% in the alternative allele (log scale). **(C)** Compares the effect sizes (beta, β) of these same BC-associated variants between the FG and MVP-EUR cohorts. The red dashed box highlights a variant with a significant difference in effect size between the two populations. Double-sided arrows indicate variants present in the FG cohort but missing from the MVP-EUR cohort. **(D)** Scatter plot of the AF between individuals in FG12 and MVP participants of African ancestry (MVP-AFR). (**E)** Scatter plot of the AF between individuals in FG12 and MVP-AFR of 10% to 0.1% in the alternative allele (log scale). **(F)** Compares the effect sizes (β) of variants between the FG12 and MVP-AFR cohorts. The red dashed boxes highlight variants with notable differences in effect size, including some that show an increased risk in the FG cohort but a protective effect in the MVP-AFR cohort (e.g., ADAMTSL1, APOBEC3A). Conversely, variants in MBD6, SETBP1, and RP11-13A2.5 show an opposite trend. This suggests potential ancestry-specific effects or differences in linkage disequilibrium (LD). A full list of all 137 associated variants and their statistical information is available in Supplementary **Table S4**.

**Fig. 4A** shows that the variant frequencies between the Finnish and other European populations are generally similar, as most data points are close to the diagonal line. This is further supported by **Fig. 4B**, which shows the same trend for rare variants. In **Fig. 4C**, we see a good agreement in the effect sizes (β) between the FG12 and MVP European-ancestry (MVP-EUR) cohorts. However, about a quarter of the variants from the FG cohort did not have a match in the MVP-EUR data, which are likely due to founder effects and population bottlenecks specific to Finland. A similar analysis was performed for the MVP African-ancestry (MVP-AFR) group **(Fig. 4D)**. Here, 29% of the variants from the FG cohort could not be matched. The data points for this comparison are more scattered, indicating a significant difference in allele frequencies (AF) between the two populations. This trend is also evident for less common variants **(Fig. 4E). Fig. 4F** displays the effect sizes (β) for the FG12 and MVP-AFR cohorts. While there is a general agreement, some variants showed marked discordance in their effect size, with some even having opposite effects (e.g., increasing risk in one population but being protective in the other). These variants, associated with genes like ADAMTSL1, APOBEC3A, MBD6, SETBP1, and RP11-13A2.5, suggest differences in LD or ancestry-specific effects. We also considered the possibility that some missing variants were due to technical or methodological inconsistencies in quality criteria, genotyping of the DNA array and imputation protocols.

We hypothesized that genes with a strong effect size (β >|0.1|) carry a stronger impact on population-specific risk variants. For instance, a missense variant in the RPA1 gene showed a strong effect in the FG12 population (β = 0.261), but a conflicting association in the MVP-EUR group **(Fig. 4C)**. RPA1 is involved in DNA repair and is functionally connected to the BRCA1/2 pathway. We confirmed that this apparent conflict was due to a small number of cases in the MVP cohort, meaning that in this case, the conflicting trend was not statistically significant.

### A pleotropic nature of BC associated variants with moderate effect size

The existence of common BC predisposition variants across multiple populations may be explained by their pleiotropic effects [49]. Comparing the FG12 cohort with the MVP African-ancestry (MVP-AFR) cohort revealed several variants with inconsistent effect sizes, sometimes with opposing directions **(Fig. 4F). Fig. 5** illustrates the pleiotropic nature of these variants, showing their representation in PheWAS and their ancestry-specific effects (Supplementary **Table S4**). Of the 137 BC-associated loci identified in the FG meta-analysis, 42% had a moderate effect size (β >|0.1|). **Fig. 5A** analyzes a specific variant, rs10995187, located next to RP11-13A2.5, which is a lncRNA gene with an unknown function. This variant has an AF of 20% in Europeans but is extremely rare in East Asian populations (AF: 1.3e-03). It shows a significant negative association with both malignant BC and benign breast conditions, suggesting it may play a protective role in various aspects of breast physiology.

**Figure 5.**
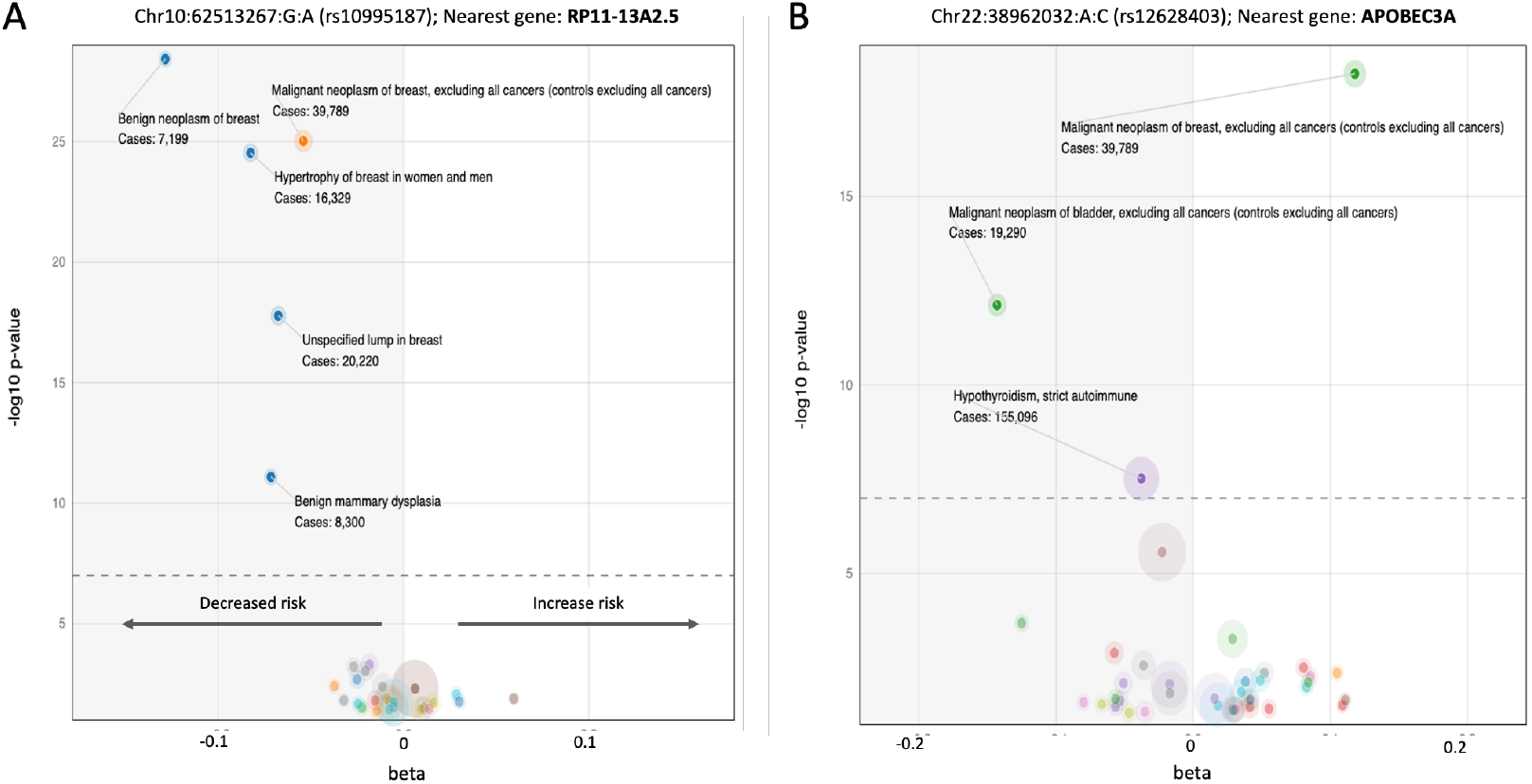
Lavaa plot representation from PheWAS for FG associated genes (FG12) with moderate effect size (β>|0.1|). Each plot visualizes a single nucleotide variant (SNV) the effect size (β, x-axis) and its direction and the significance of the association by log_10_ (P-value). The dotted horizontal line indicates the genome-wide significance threshold (p-value 5e-08). Negative beta values (left side, gray background) indicate a protective effect, or a decrease in risk. Significant phenotypes are listed along with the number of cases. Only significant phenotypes are annotated. The size of each bubble corresponds to the number of cases for that specific trait. **(A)** Chr10:123,232,597-A (rs10995187). This plot shows associations for a variant near the RP11-13A2.5 gene. **(B)** Traits association to chr22:39,599,242-C. This plot shows associations for a variant near the APOBEC3A gene. A full list of the 137 significant variants, including their ethnic distribution and associations in other cohorts, is provided in Supplemental **Table S4**.

The case of the APOBEC3A gene-associated variant further highlights this complex nature, as it shows a strong protective effect against bladder cancer but a significant increased risk for BC. The coexistence of these conflicting trends, with positive and negative beta values demonstrates that variants can have a specific, and sometimes opposing, impact on different cancer types.

The pleiotropic nature of a variant adjacent to the TTC28 gene (Chr22:28365160:C:T, rs62237617) is illustrated in Supplementary **Fig. S3**. The TTC28 gene is involved in centrosome regulation and genomic stability. This single variant is significantly associated with 10 different traits, many of which are malignancies (e.g., breast, thyroid, colon, bladder, and kidney cancers) but also include conditions related to genome stability in blood cells, such as leukemia and lymphoma. The strong pleiotropic effect of a variant affecting TTC28 gene highlights it as a potential, previously overlooked, predisposition risk gene for multiple cancer types. We conclude that examining PheWAS for pleiotropic effects is a valuable method for identifying variants according to their potential impact on many traits. It also shows that BC predisposition may impact non-coding genes (**Fig. 5A**), while variants may also exhibit opposing risk directions (**Fig. 5B**). We argue that pleiotropic variants contribute to the understanding of biological pathways, thus improving interpretability for risk prediction. Carriers of pleotropic variants in the TTC28 gene may face elevated risk for multiple cancers.

### Gene-based functional model of PWAS with inheritance mode

The PWAS is a gene-based model that aggregates the effects of each variants within the gene to assess the combined effect of damage to the protein function at an individual. The difference in the values of the gene-effect size for cases and controls determines the PWAS statistics [50]. For the group of European patients diagnosed with BC (ICD-10 G50; covering 10,682 and 138,504 female cases and controls, respectively), PWAS identified two genes based on variants affecting the coding (and splicing sites within) in females. Both genes, CHEK2 and CCDC170 were identified with a dominant inheritance mode (Supplementary **Table S5**) [36, 50]. Note that when all Europeans were included (both sexes) also TSC22D3 (ChrX, also called GILZ) was identified as significant. ICD-10 C50 is identical to the participants included in PheCode PP174.11 (Malignant neoplasm of female breast (Supplemental **Fig. S1**).

As shown in **Fig. 4**, other ethnic groups beyond Europeans increases diversity. We therefore conducted PWAS test on the global UKB (including Europeans) and used the UKB recent version (version Q3, 2022) to update the number of patients diagnozed by ICD C50. Supplementary **Table S5** shows the gene-based significant list for genes from females diagnosed with C50 (16,839 casesa; 216,529 controls), identifying 8 genes (NEK10, CCDC170, CHEK2, MRPS30, MTMR11, CRLF3, LSP1 snd FKBP5). While the PWAS method provides statistics for each gene on the assumption of a dominant, recessive or hybrid mode of inheritance [36], none of the 8 identified gene is supported by a recessive inheritance. However, when with unified number of UKB cases (16,975) within the entire population (males and females, with 410,114 controls), few more genes were identified (total 12). Among these additional 4 genes, two genes are located in ChrX (TSC22D3 and CHST7), with CHST7 and C4BPB were suported entirely by a recessive inheritance. The inheritance mode associate with any of the 12 PWAS identified genes is illustrated in Supplementary **Fig. S4**.

We confirmed these findings by testing the results from the FinnGen (FG12) dataset. Note that the FG analysis is not restricted to the coding region. We found that two of the genes were also identified as BC predisposition genes in FG12. CHEK2, a BC predisposition gene, was supported by three variants, the most significant being an intron variant (rs186430430, AF 0.8%) with an odds ratio (OR) of 2.42 (p-value 2.6e-78). CCDC170 was identified through an intergenic variant (rs6900157, AF 21.6%) with an OR of 1.1 (p-value 5.0e-16). Notably, CCDC170 was also found on the OT platform as a gene that remained after filtering for false positives (**Table 2**). However, the gene TSC22D3 was not detected in the FG12 dataset. We conclude that PWAS provides a very conservative gene list with minimal risk of false positive discovery (discussed in [50]). Moreover, it presents the value of considering gene recessive inheritance, where at least two damaging mutations affect both alleles of a gene per an individual.

### Robust BC predisposition gene list and transcriptome-based associations

We aimed to create a robust catalog of BC susceptibility genes by integrating and confirming results across different populations and study types. Instead of simply using the joint meta-analysis results from FG12, UKB, and MVP, which together included 39,789 cases and 1,002,960 controls, we focused on using a credible gene list from FG12. This approach provides a more precise variant-to-gene (V2G) mapping.

To build a consensus catalog, we first checked for consistency within the same population. We compared FG freeze 11 (FG11; 20,586 cases and 201,494 controls) to FG freeze 12 (FG12; 24,270 cases and 222,078 controls). The overlap between the gene lists from FG11 and FG12 was highly significant, with a majority (62%) also overlapping with the OT list of genes with GA score ≥0.5. This overlap was extremely significant (hypergeometric p-value 2.8E-21) **(Fig. 6A)**. We identified a core set of 20 highly confident BC predisposition genes, including 17 that were shared among all three lists and 3 additional genes from FG11 or FG12 that had strong OT support (GA ≥0.75). Note that while FG data also cover non-coding RNAs, the OT list, only consider coding genes.

**Figure 6.**
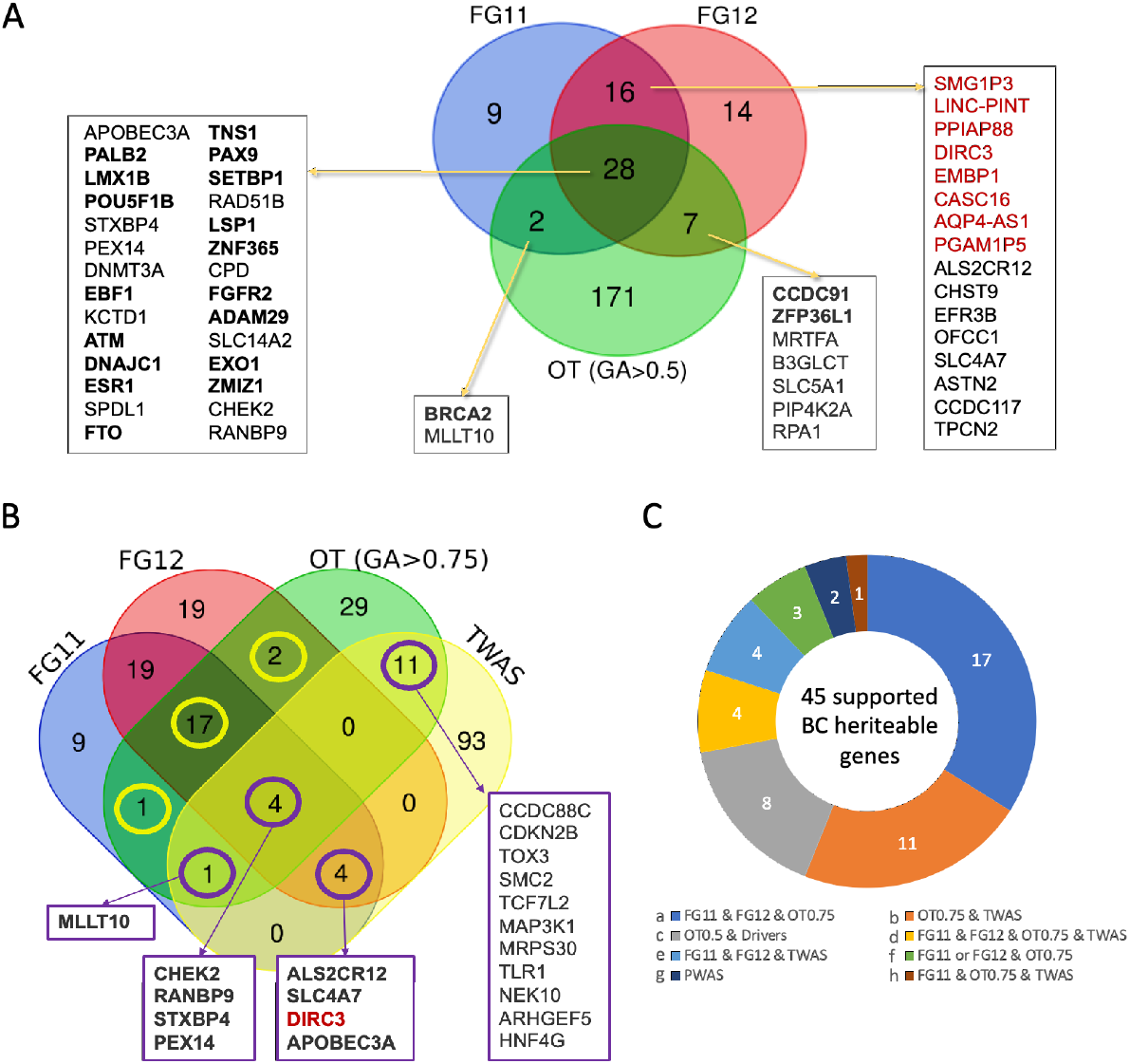
Venn diagrams of gene lists across cohorts. **(A)** FinnGen credible variants and the associated genes from freeze 11 (FG11), freeze 12 (FG12) and the list of 208 genes from the OT at GA score >0.5. Detailed summary statistics of the credible variants set (CS) for FG11 and FG12 are available in Supplementary **Table S6** and **Table S7**, respectively. Shared genes are listed, and in bold are the genes that compiled with GA score >0.75 (20 genes). Marked in red are lncRNAs that are shared between FG11 and FG12. In black are the coding genes. **(B)** Venn diagram of FG11, FG12, OT with GA score >0.75 (OT0.75) and genes associated with 79 TWAS loci. TWAS details statistics are provided in Supplementary **Table S8**. The yellow circles are the 20 genes as listed in A, and the purple circles are genes shared with TWAS results. In bold face are genes with strong evidence and are included in the core BC predisposition genes. Red color indicates lncRNA. **(C)** Pie chart of all the core BC predisposition genes by their evidence (labelled a-h). The data are based on the Venn diagram in A and B. OT at GA score >0.5 and >0.75 are labeled OT0.5 and OT0.75, respectively. The list of 50 genes with evidence represent 45 unique genes.

To expand this 20-gene core list, we applied Transcriptome-Wide Association Study (TWAS) for gene discovery. This method links genetic variants to disease by predicting gene expression from genetic data and then testing if this predicted expression is associated with the trait. TWAS is particularly useful for identifying variants in regulatory elements, which can have a strong effect on gene expression but may not be picked up by traditional methods.

We identified 79 statistically significant loci in TWAS analysis (Supplementary **Table S8**). TWAS mapping from variant to gene is even more challenging that in classical GWAS. A single locus can be associated with multiple genes (2 to 10), making a precise locus-to-gene (L2G) assignment difficult. The resulting TWAS significant finding of 79 loci (Supplementary **Table S8**) is expanded to 141 listed genes (125 unique) among them 13% are lncRNAs (16 genes). We found 20 genes from the TWAS analysis that were also supported by at least one other gene set (**Fig. 6B**). The most confident group of shared genes, found in TWAS, both FG cohorts, and the high-confidence OT set, included CHEK2, RANBP9, STXBP4, and PEX14. While CHEK2 is a known predisposition gene, the others were not previously directly associated with BC predisposition.

**Table 3** lists the genes supported by confident evidence from our analysis. This list compiles results from complementary approaches to BC genetics, including GWAS, PWAS, and TWAS, together with an assessment of evidence quality. Based on these sources (labels a–h), 45 unique genes were identified. Evidence from FG12 coding variants (p-value <5e−07) further reinforced our findings. Although some associated variants had AF <0.5%, rare variants were not explicitly included. **Table 3** shows that 15 of 30 reports of highly significant coding variants within genes listed in FG12 are included in the core BC susceptibility gene list. Of these, 11 genes have literature support for BC predisposition (binomial test, p-value 1.47e-03). This confirms that previous knowledge captured the importance of coding variants within genes. To substantiate the evidence and support for the core BC list (total 38 genes), we included as evidence the ExPheWAS significant list (q-value <5e-07, Supplementary **Table S9**) based on 17,314 cases and 192,986 female controls from UKB. The list from ExPheWAS includes 21 genes, 9 are non-coding belong to lncRNAs and another 9 supporting the findings in **Table 3**. Several genes are supported by multiple variants, and significant evidence was linked to multiple BC–related phenotypes **(Table 3)**. For example, CCDC170, identified by PWAS (evidence g; Supplementary **Table S3**), is additionally supported by FG with two variants and six breast-related phenotypes (**Table 3**). Note that 11 genes are supported only by evidence b (OT0.75 and TWAS, **Fig. 6C**) are not validated (marked with an asterisk and gray background in **Table 3**) and 7 such genes lacking any further support are to be excluded from the high confidence list. Following filteration, the BC predisposition includes 38 core genes. Notably, for about one-third of these 38 genes, there is no prior literature linking them to BC risk. Interestingly, several genes that signify subtypes of BC (e.g., ER positive or ER negative,Supplementary **Table S10**) are included in the core set of 38 predisposition genes (**Table 3**, Supplementary **Fig. S5**). We tested the current presence of clinical panels and marked 4 of the genes that are already included in commonly used Invitae and Myriad MyRisk hereditary BC panels **(Table 3)**. Inspecting the functional annotation of the BC predisposition core set emphasize the importance of genes acting in DNA repair, cell cycle, adhesion, hormonal regulation, epigenetics and transcription regulation.

**Table 3.**
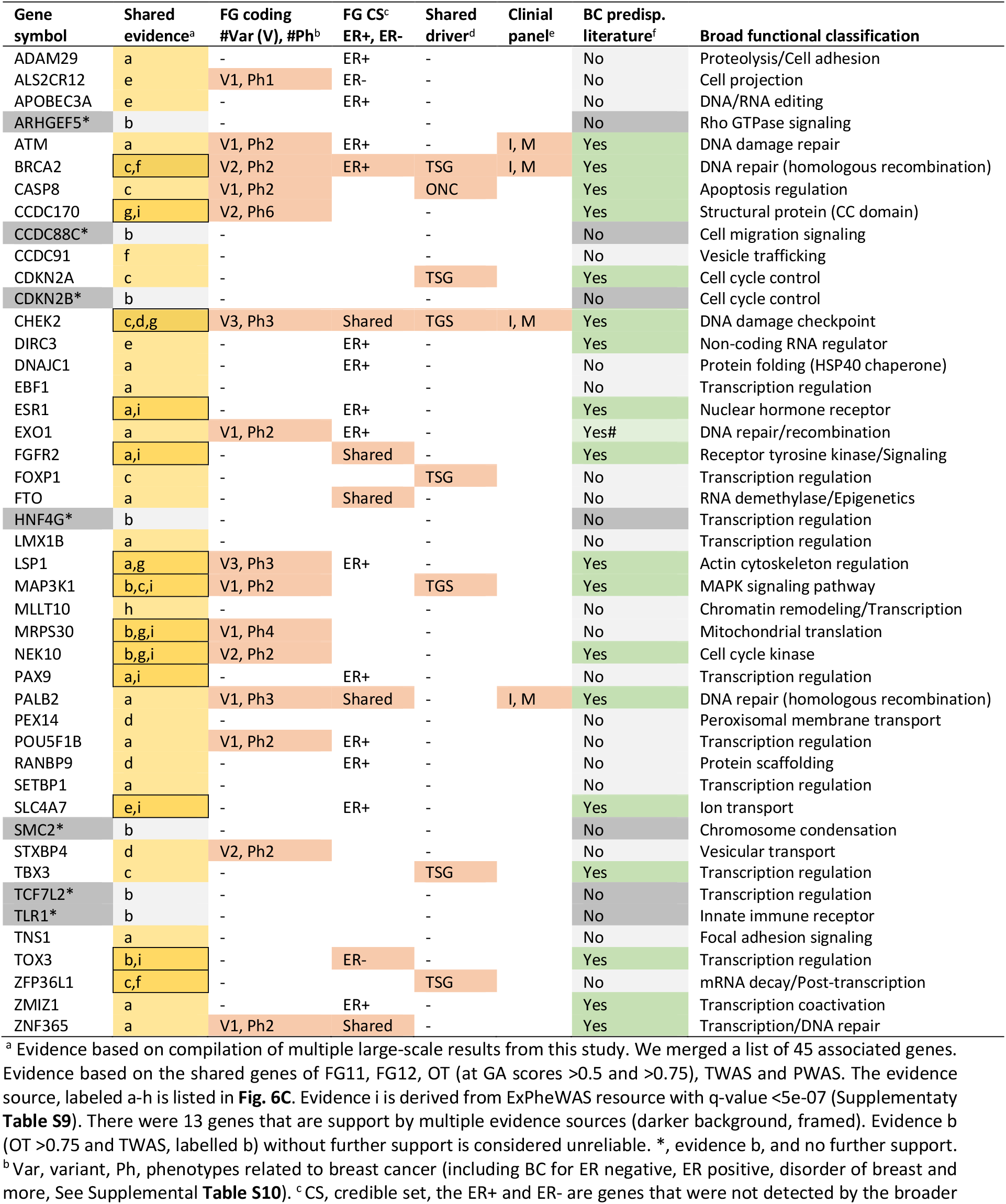
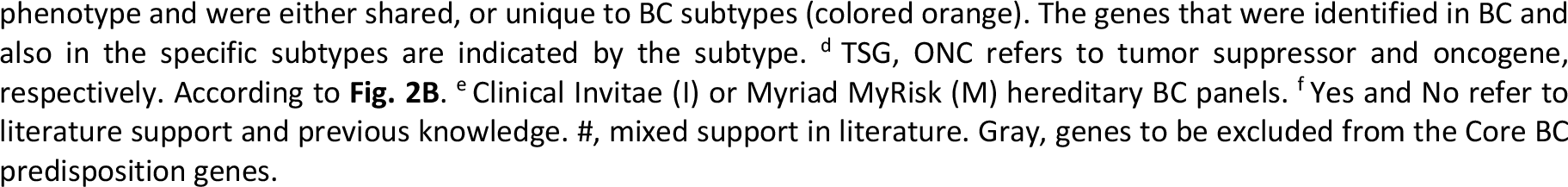
List of 45 candidate genes (included 38 core set) identified by a unification of results from large-scale association studies (GWAS, TWAS and PWAS)

### Gene-based PWAS for population stratification of BC predisposition

To further interpret the relevance of identified genes (as in PWAS), we illustrate the utility of the associations in view of the population **(Fig. 7)**.

**Figure 7.**
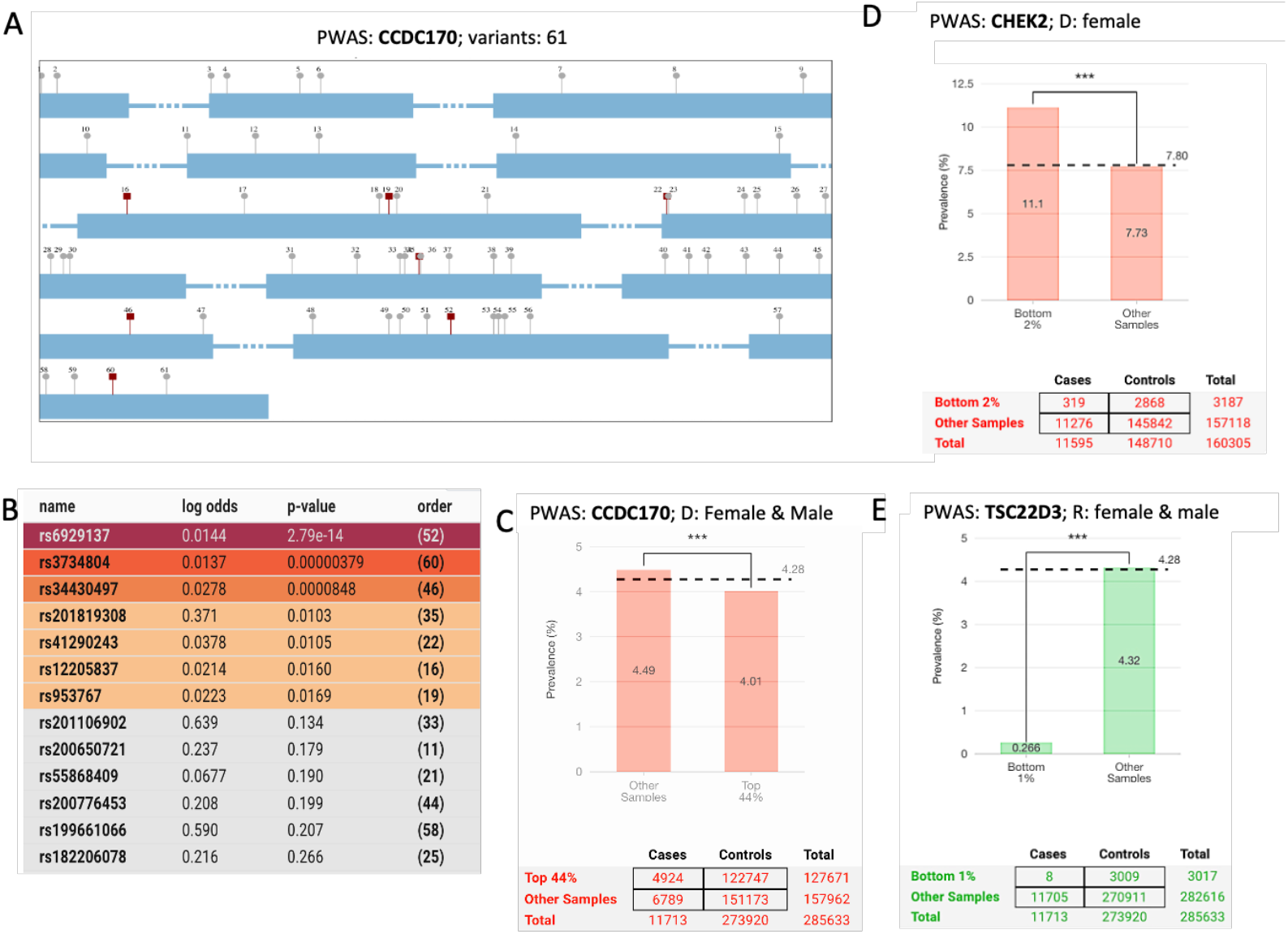
Population-level interpretation of significant PWAS associations in BC. **(A)** Schematic of the coding region of the CCDC170 genes and its exon partition. The 7 nominally significant variants are indicated in red rectangles. **(B)** Among the 61 coding variants, rs6929137 reached the genome-wide significance threshold, while the other 6 variants are colored according to their log-odds values (see [36]). **(C)** Population partitioning and BC prevalence. “D” and “R” refer to dominant and recessive models, respectively. The top 44% of individuals had a slightly reduced prevalence (4.01%) compared to the remainder (4.49%), with the baseline prevalence indicated by the horizontal dashed line (4.28%). *** indicates statistical significance (<5e-4) of the population partition. **(D)** CHEK2 dominant effect scores highlight a high-risk subset. The bottom 2% (319 individuals) had a BC prevalence of 11.1% compared to 7.8% overall (females only). **(E)** TSC22D3 recessive (green) effect scores identify a subgroup with reduced risk. Specifically, the bottom 1% (8 individuals) had a markedly lower BC prevalence (0.27% vs. 4.28% overall). The number of individuals in each partition for case, controls, and total **(C-E)** is shown.

**Fig. 7A** depicts the scheme of CCDC170 and the position of all 61 variants within its coding region. Among these, 7 of 61 variants are marked in red as significant (p-value <0.05; **Fig. 7B**). However, only a single variant (rs6929137, variant 52) reached genome-wide statistical significance. **Fig. 7C** demonstrates how BC risk changes as a function of PWAS effect scores in the UKB cohort. Recall that lower PWAS effect scores indicate greater gene damage [34]. The bar plots illustrate the most significant partitioning of individuals by dominant effect scores and the corresponding fraction of BC cases. For example, in the association between BC and dominant effect scores of CCDC170, a dashed line shows the baseline BC risk across the cohort (4.28% cases in both sexes). The bar marked as “Top 44%” refers to the 44% of the cohort with the highest PWAS dominant effect scores of CCDC170, with an estimated BC prevalence of ∼4.01%. This translates to 4,942 individuals with the highest dominant effect scores who had BC, compared to a higher prevalence (4.49%) in the rest of the cohort. **Fig. 7D** shows population partitioning by PWAS effect scores for CHEK2 under the dominant model. Here, the effect score reflects whether an individual carries at least one damaging variant. The bottom 2% (319 cases) represent the best-partitioned group, where BC risk increases to 11.1% (compared with 7.8% on average; females only). **Fig. 7E** shows population partitioning by PWAS effect scores for TSC22D3 (ChrX, also called GILZ) under the recessive model, where the score reflects the presence of at least two damaging alleles. The bottom 1% represents the best-partitioned group (males and females). For 8 patients in this group, the damaged gene led to a markedly reduced BC risk (0.27% compared with 4.28%). This supports TSC22D3 as carrying protective variants for BC risk in a small subset of individuals under a recessive model (**Fig. 7E**). This gene was also identified as significant by conducting PWAS with the entire population (including non-Europeans; Supplemental **Table S5**).

We showed that gene-based partitioning of the population is clinically valuable, as it highlights individuals with extreme effects (either increased or protective risk) and quantifies the number of diagnosed patients who may benefit from gene-targeted therapies. It also allows exploration of recessive inheritance patterns in both families and populations, an aspect largely overlooked by classical GWAS approaches.

## Discussion

This study revisits current knowledge on BC predisposition genes and proposes a protocol for ranking strongly supported BC susceptibility genes while minimizing false positives and weakly supported candidates. A major source of false positives arises from the inconclusive mapping of lead SNPs to genes. In gene-dense loci, linkage disequilibrium (LD) often results in the assignment of dozens of genes to a single associated variant [51]. In this study, we chose to address the gene (and not specifically the leading variants). In traditional GWAS, the associated variant is reported at the nearest gene, which is an oversimplification and ignores complex genomic architecture. Even when chromatin interactions are considered, regions with poorly annotated genomic structures often suffer from incorrect gene assignments [52, 53]. While GWAS has uncovered hundreds of common risk loci for BC, these variants individually confer only modest increases in risk and cumulatively explain a limited fraction of heritability [54]. Despite advances in assigning statistically significant variants to likely causal genes through fine mapping and co-localization approaches [55], much of the inherited risk of BC remains unexplained. Quantifying the proportion of BC heritability explained by common variants emphasizes this gap [56]. A systematic study of conservative BC genes identified 15 new loci explaining approximately 2% of familial risk [28]. In other studies, it was estimated that low penetrance loci together explain <4% of the familial relative risk [18]. In total, more than 80 known loci account for roughly 16-18% of familial risk [28, 57]. In the present work, we incorporated complementary approaches from GWAS, TWAS, and PWAS. We emphasise coherence across different sources rather than merging them into a single cohort. While this strategy reduces statistical power, it enhances consistency, as meta-analytic signals can be obscured by founder effects or technical limitations in mapping and imputation [58, 59].

By maintaining a gene-centric perspective, we argue that the conservative framework adopted here enables more robust mechanistic interpretation. The foundation of PWAS is the impact of mutations on the function of the encoded protein. Therefore, it is resistant to mapping issues that strongly affect other association methods (e.g., TWAS and GWAS). Notably, including changes in gene expression data as a proxy for genetic signal through TWAS neither contributed to nor altered the risk associations of candidate genes, consistent with observations from a large study of more than 46,000 women with BC [60]. Instead, PWAS is based on a very conservative approach with a small number of gene discoveries [36]. Nevertheless, the listed genes often lead to functional interpretation of biological mechanisms. This helps identify causal genes and functional pathways more accurately. Another advantage of gene-based approaches is the ability to capture compound heterozygosity and recessive inheritance patterns, which are typically missed by GWAS. Of the PWAS-identified genes, recessive inheritance is exposed. Recent PWAS findings suggest that compound heterozygosity at multiple DNA repair genes plays a larger role in BC predisposition than previously recognised [50].

Another factor that can weaken genetic signals is the inconsistency in the definition of the BC phenotype. Statistical outcomes are sensitive to cohort composition; in the case of BC, it also matters if males are included in the cohort, differences in average age across biobanks, and if the accuracy of clinical records is not always standardised. In the FG dataset used throughout this study, BC patients afflicted with other cancer types were excluded, whereas for the TWAS analyses, self-reported BC data were used. Consequently, our core BC predisposition gene set **(Table 3)** does not include broadly tumor-associated genes such as TP53, PTEN, CDH1, and STK11, which have been implicated across multiple cancer types and are confirmed to contribute to BC predisposition risk [61]. Because we seek overlap between methods and resources, and used FG cohorts that were depleted of any other cancers **(Fig. 6, Table 3)**, some classical predisposition genes that play a central role in pan-cancer predisposition were not identified (e.g., TP53, PTEN) [62].

Expanding the catalogue of BC predisposition genes according to clinical panels is critical for genetic counselling, follow-up, and targeted family screening [29]. In this study, we focus on BC cases in a broad case-control framework. We ignore the clinical category, staging, or the molecular property of the BC-affected women. Testing panels of high- and moderate-risk genes in more than 60,000 affected women revealed substantial differences in clinical subtypes and associated genetic risk [63]. For example, the highest risk for ER-positive BC is associated with ATM, CDH1, and CHEK2, whereas BARD1, BRCA1, BRCA2, PALB2, RAD51C, and RAD51D confer a higher risk for ER-negative BC [64]. Rare pathogenic coding variants in these genes further contribute to overall BC risk [12, 63].

In addition, there are more specialised cohorts of BC survivors that develop secondary primary cancers [65], BC patients that were exposed to radiation [66], and more. Such stratification is limited using population studies, as in retrospective biobanks. The contribution of rare variants from exome sequencing was not explicitly addressed in this study. As expected, rare variants are often below statistical significance [67]. Gene-based collapsing models for rare variants in UKB validated the significance of a small set of high BC risk (BRCA1, BRCA2, PTEN, PALB2) [63]. Recent large-scale analyses testing the impact of protein-truncating variants (LoF mutations) confirmed the known high-penetrance genes (ATM, BRCA1, BRCA2, CHEK2, PALB2), with only MAP3K1 reaching exome-wide significance (**Table 3**). Other candidates, including CDKN2A and BARD1, did not reach statistical significance.

In this study, we have not explicitly analyzed the contribution of rare variants to candidate genes. Overall, the rare variants’ impact on coding pathogenic variants appears modest, suggesting that even the combination of rare and common variants leaves a substantial fraction of genetic susceptibility unexplained [68]. While rare variants (<0.01% in the population) may have a limited contribution to screening efforts, they can benefit selected sub-populations. For example, RECQL is a DNA helicase that acts in preventing double-strand breaks. RECQL was recognized as a new BC susceptibility gene among Polish and Quebec populations, where each population is signified by a unique rare loss of function (LoF) variant [69]. Other examples of rare and ultra-rare LoF variants substantiate the importance of DNA repair mechanisms in the predisposition to BC [70]. Moreover, among BC-affected families from restricted populations, rare missense variants expose understudied cellular mechanisms that increase BC risk [45].

There are a few directions that can advance the clinical relevance of our findings. Our unified pipeline narrows the BC predisposition landscape to 38 high-confidence genes, refer to core BC gene set **(Table 3**). Eleven of the core BC genes already carry convincing epidemiological evidence for BC risk (binomial enrichment p = 1.47e-03), among them four (ATM, BRCA2, CHEK2, PALB2) are currently interrogated by hereditary BC panels (Invitae, Myriad MyRisk, **Table 3**). The remaining genes represent actionable candidates for panel expansion, especially with supportive orthogonal validation (e.g., rare-variant, burden tests). The incorporation of genetic refined signal to the polygenic risk scores (PRS) can lead to improved personalized treatment on the BC field. Based on a large collection of studies, >170 common susceptibility loci were compiled whose individual effects are small but the combined PRS showed its value in stratifying women into subtype-specific scores. The reported PRS [56] showed that the higher 1% women at risk displayed 4.4-and 2.8-fold higher risk relative to the average women for ER-positive and ER-negative BC, respectively. We propose that incorporating the additional variants associated with the understudied core gene genes is likely to improve clinical utility and further boost the PRS. We showed that even a single gene partition of the population can benefit the risk assessment (**Fig. 7**).

The initial PWAS analysis restricted to Caucasians [36] yielded only three significant genes. Yet the contribution of high- and medium-penetrance loci to breast-cancer risk varies markedly by ancestry: ATM shows limited relevance in Asians, founder BRCA2 mutations greatly elevate risk in Ashkenazi Jews, pathogenic BRCA2 and PALB2 variants are more common in Black women, and CHEK2 frequencies are lower in both Black and Asian populations. Expanding the cohort to all UKB participants and updating case numbers from 10.6 k to 17.0 k increased the discovery to eight significant genes, five of which are included in the BC core list with multiple lines of evidence (**Table 3**). PWAS that was expanded and included non-European population identified several new genes in female group (MTMR11, CRLF3, FKBP5). The additional genes that uncovered recessive associations (TSC22D3, CHST7, C4BPB) were previously unreported for familial BC, offering promising new candidates for clinical follow-up (Supplementary **Fig. S4**). Notably, TSC22D3 and CHST7 are in ChrX, and multiple evidence connects TSC22D3 to sex-dependent genetics [71].

This study carries several inherent limitations. First, collapsing methods such as PWAS and OT are restricted to protein-coding genes, so the contribution of non-coding RNAs, especially lncRNAs, to BC predisposition remains largely unexplored [72, 73]. Second, cohort definitions were inconsistent. Specifically, the FG excluded participants with any non-breast malignancy (Supplementary **Fig. S2**), whereas TWAS and PWAS did not. Some GWAS that contributed to OT collection relied on self-reporting. Another issue is that for UKB females datafield such as ICD-10 Z85.3 (“personal history of breast cancer”) may be wrongly added to the control group, therefore the genetic might be diluted. These differences in ascertainment (i.e., cancer registry, ICD-10, PheCode, self-report) likely attenuated risk estimates. Finally, the sample was overwhelmingly Caucasian (Finns, other Europeans), with only modest numbers of Asian participants from MVP or UKB. Incorporating emerging biobanks such as All of Us and specialized national data from Mexico, Japan and China will broaden the allelic spectrum, refine risk predictions, and provide evolutionary insight into BC susceptibility.

## Conclusions

This study refines the landscape of heritable B predisposition by applying a conservative, gene-centric framework that reduces false positives and emphasizes coherence across GWAS, TWAS, and PWAS. We defined a core set of 38 high-confidence susceptibility genes, including both established drivers and novel candidates, with several supported by recessive inheritance patterns. While limited by cohort composition and a focus on protein-coding genes, our framework improves mechanistic interpretability and highlights clinically actionable candidates for panel expansion. These results highlight the value of integrating gene-based models into genetic counseling, thereby enhancing precision prevention strategies and informing targeted surveillance, early detection, and therapeutic decision-making in breast cancer care.

## Data Availability

The analysis shown in this study are supported by Supplementary materials (Fig. S1-S5 and Supplementary Tables S1-S10). GWAS (with 10 covariates) and PWAS (with 10 or 172 covariates) performed in this study for ICD10: C50 (for female only, and both sexes). The PWAS is available in https://github.com/nadavbra/pwas. PWAS analysis on Europeans is presented in PWAS Hub (https://pwas.huji.ac.il/).

## List of abbreviations

BC: breast cancer
CS: credible set
eQTL: expression quantitative trait locus
FG: FinnGen
GWAS: genome wide association study
ICD10: International Statistical Classification of Diseases and Related Health Problems,10th Revision
LD: linkage disequilibrium
LOF: loss of function
lncRNA: long non-coding RNA
MAF: minor allele frequency
MVP: million veteran program
OR: odds ratio
OT: open targets
PheWAS: phenome wide association study
PRS: polygenic risk score
PWAS: proteome wide association study
TWAS: transcriptome wide association study
UKB: UK biobank

## Acknowledgements

The authors are thankful to the CSE system for their computational support and for maintenance of the PWAS Hub website.

## Funding

This work was partially supported by the MOST project on health of the heart, grant number 2753/20 (ML), and the NAAF 2023. The work was initiated based on a grant from HUJI-Shaare Zedek Medical Center(SHEMESH, 2023) on familial breast cancer.

## Contributions

Study conception: ML Data acquisition: SS, RZ. Investigators, ML, RZ, SS, AS. Data analysis: ML, RZ, SS, AS. Creation of new software used in the work: RZ, ML. Drafting the work: ML Approval of the submitted version: all authors. Agreement on both to be personally accountable for the author’s own contributions: all authors. All authors read and approved the final manuscript.

## Ethics declarations

The study was approved by the University Committee for the Use of Human IRB ethical approval and written informed consent were obtained by The Hebrew University. Research Approval number 12072022 (July 2025). This study uses the UK-Biobank (UKB) application ID 26664 (Linial lab).

## Consent for publication

Not applicable.

## Competing interests

The authors declare that they have no competing interests.

## Supplementary Information

Tables S1-S10

Figures S1-s5.

## Notes

### Competing Interest Statement

The authors have declared no competing interest.

