## Supplementary material for "Integrative Gene-Centric Analysis of Breast Cancer: High-Confidence Germline Predisposition Genes from Population-Scale Cohorts": Figures S1-S5

**Supplementary Tables**

Supplemental **Table S1:** Open Targets (OT) list of GA scores along with supporting evidence.

Supplemental **Table S2:** Classical GWAS for ICD10 C50 (PheCode 117.4) using all UKB for female cohort.

Supplemental **Table S3.** Coding gene GWAS (cgGWAS) from all females in the UKB cohort, p-value <1e-05.

Supplemental **Table S4**: FG (freeze 12) for BC using meta-analysis with 137 associated loci.

Supplemental **Table S5**: PWAS results for BC PheCode 174.11 / ICD-10: C50 Europeans and Global

Supplemental **Table S6:** List of C3_Breast_EXALLC from FG freeze 11 (FG11) creadible set (CS).

Supplemental **Table S7:** List of C3_Breast_EXALLC from FG freeze 12 (FG12) creadible set (CS).

Supplemental **Table S8:** List of 79 loci from TWAS and associated statistics based on GWAS eQTLs.

Supplemental **Table S9:** List of EXPheWAS (174.11) with corrected p-values for multiple phenotypes and genes. Supplemental **Table S10:** Summary statistics of C3_Breast_(ERPLUS and ERNEG)_EXALLC from FG12.

**Supplementary Figures**

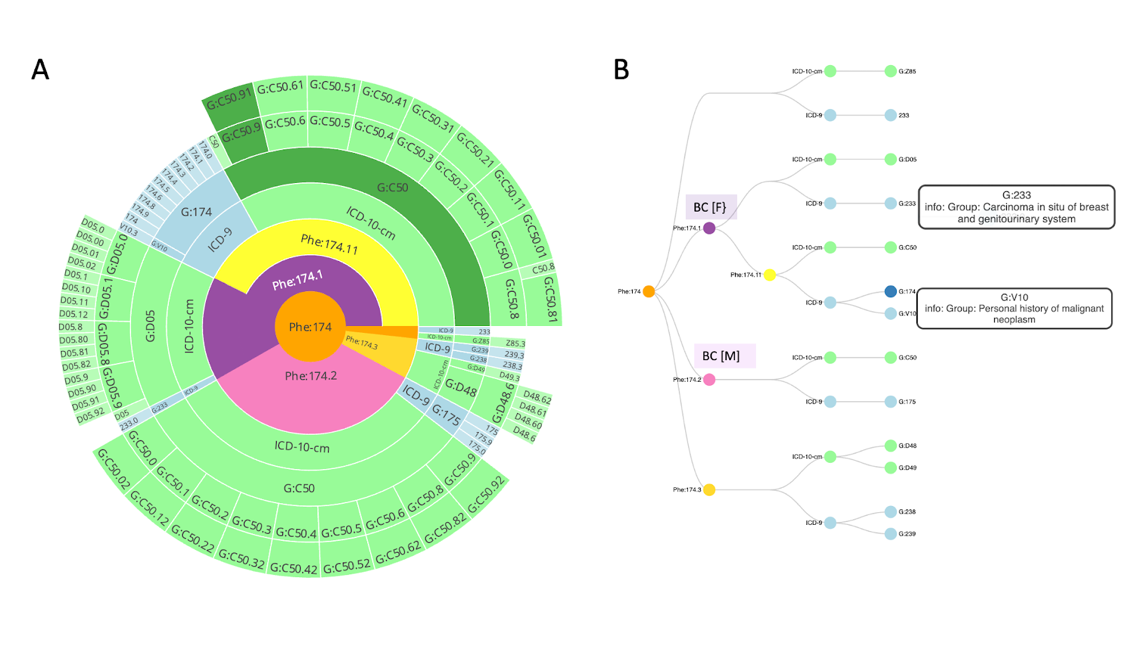

**Figure S1.** Mapping of PheCode with respect to ICD-10 (and ICD-9). **(A)** Circular presentation of PP174.11 that match C50 in ICD-10. The analysis performed on pheCode 174.11 (yellow) that covers the ICD-10 C50 (dark green). The G:C50 signs that the group includes children codes (shown in an external circles). (B) The same analysis according to a tree-like presentation. We focus on breast cancer of females (BC [F]) and had not included the BC of males (174.2). We show full names some of the subtree labels for ICD-9.

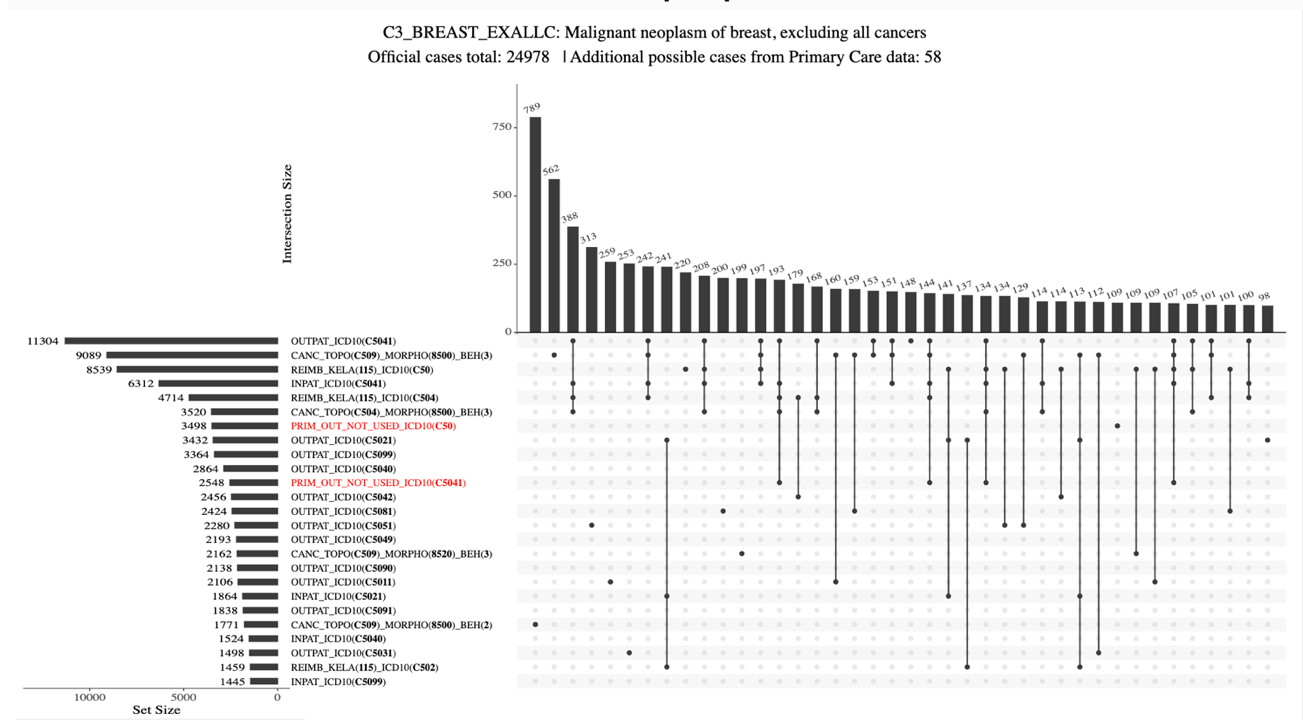

**Figure S2.** Upset plot for FinnGen (FG12) for BC with 24,978 individuals. The plot lists the the number of cases (with intersection) when combining different types of data source for breast cancer. The plot also marks the excluding sets of all other cancer. The BC phenotype title is shown. The labels on the left refer to the datasets used (and the number of individuals). The lines and dots show which datasets contribute to each intersection

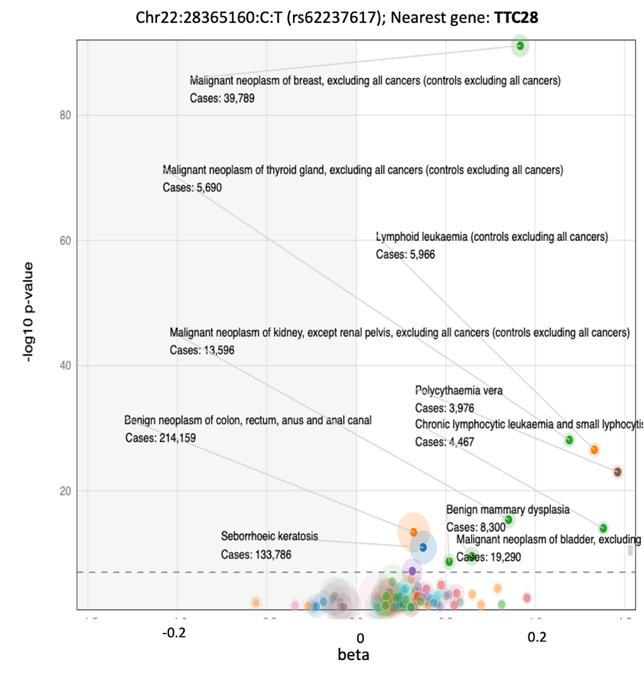

**Figure S3.** Lavaa representation for Chr22:43810601-C. The nearest gene is TTC28. The dotted horizontal line indicates the genome-wide significance threshold (p-value 5e-08). Negative beta values (left side, gray background) indicate a protective effect, or a decrease in risk. Significant phenotypes are listed along with the number of cases. Only significant phenotypes are labelled. The size of each bubble corresponds to the number of cases for the specific trait. This variant is associated with an increased risk for BC, thyroid cancer, kidney cancer, and hematologic malignancies. The multiple significant associations with increased risk imply a pleiotropic effect on malignancies.

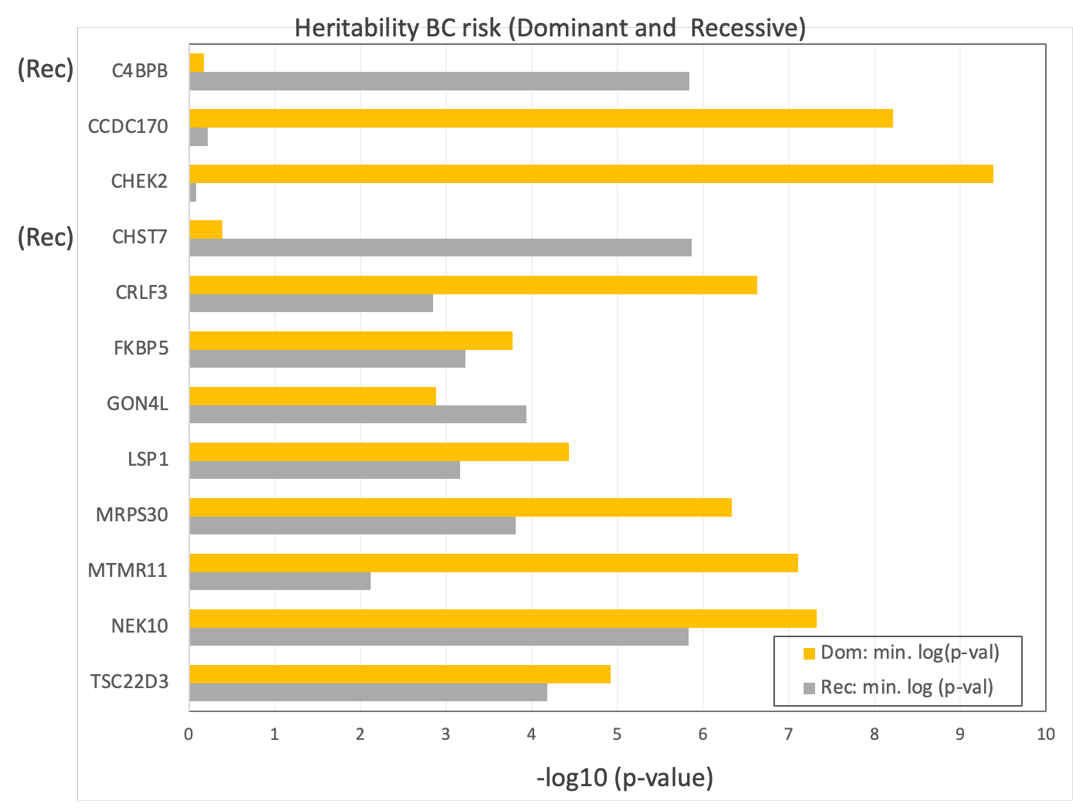

**Figure S4.** Histogram of the significant association signals (with FDR <0.05) for candidate BC ICD10:C50 from both females and males for susceptibility genes. Bars show the–log10(p-value) observed for each gene under either dominant (yellow) or recessive (gray) models. Genes surpassing this threshold are labeled and ordered alphabetically. Rec, genes that signified by a recessive inheritance.

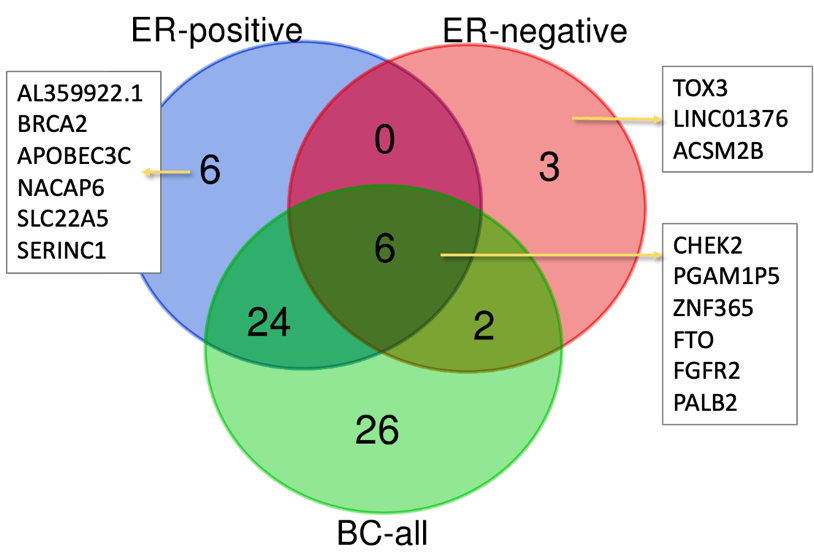

**Figure S5.** Venn diagram of the FinnGen credible variants set (CS) by their gene symbols from three cohorts in FG12. All variants that failed mapping (marked as N.A.) were removed from the analysis. The list covers the BC-all (64 mapped, 58 unique genes). Another 11 and 39 (36 unique) genes derived from ER negative and ER positive cohorts, respectively. The genes that are shared and those that specify the BC subtypes are listed. The gene list is available in Supplemental **Table S10**.
